# The Interplay Between Sex, Rater, and Autism Polygenic Scores in Autistic Traits in General Population Adolescents

**DOI:** 10.64898/2026.11.17.26367610

**Authors:** Thomas Sollie, Melanie M. de Wit, René Pool, the Autism Working Group of the Psychiatric Genomics Consortium, Meike Bartels, Tinca J.C. Polderman, Wonuola A. Akingbuwa

## Abstract

**Background:** Autism is diagnosed more often in males than in females, but it remains unclear to what extent this reflects sex differences in autistic traits themselves or in how those traits are recognized. We compared up to four raters of the same participants, testing whether autistic trait scores differ by sex across raters, and whether the association with autism polygenic scores (PGS) differs by sex (genetic sex amplification) or rater.

**Methods:** We derived autistic trait scores from mother- (*n* = 4,541), father- (*n* = 3,411), teacher- (*n* = 2,939), and self-reports (*n* = 2,632) on an autistic traits subscale of the Achenbach System of Empirically Based Assessment for 12–14-year-olds in the Young Netherlands Twin Register (total *n* = 5,965). Ordinal mixed-effects regression modeled a three-way interaction between PGS, rater, and sex. Measurement invariance analysis tested the subscale items for sex bias.

**Results:** Mothers, fathers, and teachers rated males higher than females (odds ratios (*OR*) = 1.56, 1.49, and 2.34), whereas females rated themselves higher than males did (*OR* = 0.71, males as reference). Autism PGS were associated with higher autistic trait scores (*OR* = 1.22), but this association did not differ by rater or sex. One item (*“Acts too young for his/her age”*) was biased toward males in mother- and self-ratings.

**Conclusions:** These rater effects are consistent with under-recognition of autistic traits in females by external raters. In contrast, the PGS association did not differ by sex or rater, providing no evidence for sex amplification, though current autism PGS have limited predictive power.

## Introduction

Autism is a heterogeneous neurodevelopmental condition characterised by differences in social communication, sensory processing, and restricted/repetitive behaviours^1^. It shows a male to female diagnostic ratio of 4 to 1^2,3^. This raises the question of whether the diagnostic sex ratio reflects differences in autistic trait levels^4^, or in how the traits are recognised by others, such as via rater effects^5^. Most research has used primarily male samples, prompting increasing investigations of sex differences in autism^6,7^. In this study we investigated the interplay between genetic and rater effects on sex differences in autistic traits.

Autism is highly heritable, with estimates of around 70–90%^8,9^. Genetic effects can be investigated using polygenic scores (PGS)^10,11^: a weighted sum of an individual’s trait-associated alleles, with weights based on population-level estimates derived from genome-wide association studies (GWAS)^10^. Sex differences have been reported in the association between the autism PGS and specific autistic traits^12–14^. Furthermore, our previous work found that autistic females (without intellectual disability) have a higher mean autism PGS than autistic males^14^, but whether this reflects differences in trait levels or in external recognition remains unclear. One way in which genetic factors could contribute to the diagnostic sex ratio is sex amplification, i.e. the same autism-associated variants having systematically larger effects in one sex than in the other^15^. Larger effects in males would be consistent with the higher heritability of autism reported in males^8^. Under sex amplification, the PGS would be more strongly associated with autistic trait levels in males, so that at high PGS, males would have higher trait levels than females with the same PGS, and more males would exceed the diagnostic threshold. Sex amplification can therefore be tested directly as a sex difference in the association between the autism PGS and autistic traits.

Autistic trait recognition is important for receiving an autism diagnosis. Whether a child or adolescent is referred for assessment largely depends on external observers, typically parents and teachers^16,17^. Importantly, different raters agree only moderately on autistic traits^18–20^, and traits in females may be less readily recognised by external raters^19,21,22^.

Finally, although PGS have been used to study sex differences in autism and autistic traits^14^, many studies implicitly assume that different raters report the same underlying autistic traits, so that genetic associations do not depend on the rater. Riglin et al.^23^ compared PGS associations between parent- and self-reported autistic traits in young adults, but used a different questionnaire for each rater, did not formally test whether associations differed between raters, and examined sex only in stratified sensitivity analyses. If parents, teachers, and individuals themselves capture different aspects of autistic traits, genetic associations may also differ between raters. Conversely, if sex amplification acts on autistic traits themselves, a sex difference in association should appear regardless of who reports the traits.

In this research, we derived up to four autistic trait scores for each participant from mother-, father-, teacher-, and self-reports, using the same measure across raters. We regressed the scores on autism PGS, participant sex, and rater, including their two- and three-way interactions. This model allowed us to address four research questions. First, we investigated genetic sex amplification by testing whether the association between autism PGS and autistic trait scores was moderated by sex (PGS × sex). Second, we tested the interaction between rater and sex (rater × sex) to address autistic trait recognition. Third, we tested whether rater moderated the association of the PGS with autistic trait scores (PGS × rater), to determine whether raters capture distinct genetic signal. Finally, we tested whether the PGS association differs across raters and sexes jointly (PGS × rater × sex), to determine consistency of a sex amplification effect across raters. We used an adolescent general population sample, which avoids ascertainment on clinical diagnosis and its skewed sex ratio. We tested the questionnaire items for sex bias via measurement invariance analysis. This research informs sources of bias in autism identification and phenotyping strategies in genetic research with the aim of aiding a better understanding of autism and autistic traits in both sexes.

## Methods

### On language usage

In this study, *sex*, *male,* and *female* refer to biological, chromosomal sex. We use *autism* rather than *autism spectrum disorder*, and *traits* rather than *symptoms*^24^. We use both identity-first and person-first language when referring to autistic people, reflecting their diverse preferences^25^. The term *liability* in this study refers to a statistical concept used in genetic research and is not a value statement about autism.

### Preregistration

We preregistered this study (DOI: 10.17605/OSF.IO/4PQEB) after data access but before statistical modelling. We performed exploratory analyses of the data distributions to determine the appropriate statistical model. Deviations from the preregistration are described in the Supplementary text.

### Software

We performed coding and visualisation in R version 4.5.2^26^. We performed MI analyses using Mplus Version 8^27^.

### Participants and procedure

Data were obtained from the Netherlands Twin Register (NTR)^28^, a national register that includes genomic and questionnaire data. We used data from the Young Netherlands Twin Register (YNTR) cohorts^29^, which includes the Dutch Achenbach System of Empirically Based Assessment (ASEBA)^30^ questionnaires completed by parent and teacher of children up to age 12, and self-reports from age 14. Participant sex was determined from sex chromosomes. Ethical procedures are outlined in the Supplementary text.

### ASEBA-based autistic trait scores

We used data from the following ASEBA questionnaires: the Child Behavioural Checklist (CBCL), Teacher’s Report Form (TRF), and Youth Self Report (YSR)^30^. In this study, we included participants (*n* = 6163) around 12 years of age (assessment date 1999–2022) with CBCL (mother and father rated) and TRF questionnaire data. For the YSR, we used data at age 14 (assessment date 2005–2013), the closest age to the CBCL and TRF assessments at which self-report data was available.

We derived autistic trait scores from an ASEBA autistic traits subscale validated by So et al.^31^. It consists of 10 CBCL/TRF items listed in Supplementary text. The YSR includes the same items except item 80, and was not validated in So et al.^31^.

ASEBA items are measured on a 3-point Likert scale with response options 0 = not true, 1 = somewhat or sometimes true, and 2 = very true or often true. The same scale can be calculated for the CBCL, TRF, and YSR by summing the item scores (resulting in a range of 0–20, or 0–18 for YSR). As such, autistic trait scores were available for each participant from up to a maximum of four raters: mother, father, teacher, and self.

### Autism polygenic scores

We included only YNTR participants with genotype data, and calculated autism PGS using a 2026 freeze of the case-control autism discovery GWAS of the Psychiatric Genomics Consortium^32^, which did not include (Y)NTR participants. We limited analyses to individuals of statistically inferred European genetic ancestry.

We estimated Single Nucleotide Polymorphisms (SNPs) weights from the summary statistics with *SBayesRC* in *gctb*^33,34^, and calculated PGS in *PLINK2*^35,36^. For full details on genotyping, quality control, and PGS calculation, see Supplementary text.

### Covariates

We included participant age at assessment as a covariate, as age influences CBCL outcomes and external raters’ expectations of behaviour^37^. As the social perception of autism has changed over time^38,39^, we included the questionnaire completion date (assessment date). We included genotyping platform to control for batch effects. Finally, to correct for population stratification, we included 10 genetic principal components (PCs)^40^ in the analyses^41^.

### Data cleaning and variable coding

We excluded ratings with three or more missing items. We removed participants with discordance between chromosomal sex and their reported gender as we lacked power to investigate this subgroup.

We excluded ratings with outlying ages using Tukey’s fences (1.5 x interquartile range; Supplementary text)^42^.

We scaled continuous variables including PGS, and the 10 PCs to a mean of 0 and *SD* of 1. Age and assessment date were expressed in decimal years and years respectively and mean centred across the entire sample.

The categorical variables sex, rater, and genotyping platform were effect-coded so that main effects reflect averages across the levels of the other variables rather than within a reference category.

### Statistical modelling: ordinal regression

Simulation modelling concluded that ordinal regression was more suitable than linear regression for the skewed distribution of the autistic trait score (Supplementary text and Figure S1). For this, we ordered the scores categorically into three levels: *no* (those with a score of 0), *low* (1–3), and *high* (4+). The *high* cut-off is half of the 8+ cut-off that best predicted diagnosed from combined mother-CBCL and TRF scores in So et al.^31^, as we analysed raters separately.

We ran cumulative link mixed effect ordinal models (CLMM) using the *ordinal* package^43^. CLMMs modelled an unobserved continuous variable underlying the observed autistic trait score levels, divided into three levels by two thresholds. Model estimates are expressed on this latent scale of the score: coefficients (*β*) represent log-odds ratios of belonging to a higher score level (*no* vs. *low* or *high*; *no* or *low* vs. *high*). We exponentiated coefficients into odds ratios (*OR* = *e^β^*), reported with 95% confidence intervals (CIs). *ORs* represent the change in odds per *SD* increase for PGS and principal components, per year for age and assessment date, and relative to the grand mean for effect-coded categorical variables.

Observations in our data were not independent. Ratings were nested within participants (up to four raters each), and participants within families. We accounted for this clustering with random effects for family and participant.

We ran five hierarchical models, each one expanding on the previous, which are shown in **Equation 1**. Model 1 included the covariates (genotyping platform, age of the rated individual, assessment date, and 10 PCs). Model 2 added the main effects autism PGS, participant sex, and rater. Model 3 added two-way interactions between the main effects. Model 4 added the three-way interaction. Model 5 added covariate interactions with all main and two-way interaction terms, as recommended for gene-environment interaction studies^44^. Given its complexity, we treated it as a sensitivity analysis.

**Equation 1: Hierarchical modelling formula.** Autistic trait score is modelled as a latent continuous variable underlying rating *i* of participant *j* in family *k*, using ordinal regression. Subscripts show the level at which each variable varies: rater, age and assessment date vary between ratings (*ijk*); PGS, sex, genotyping platform and PCs vary only between participants (*jk*). Random effects *u* and *v* for family and participant respectively. Two thresholds divided the latent score into three levels.

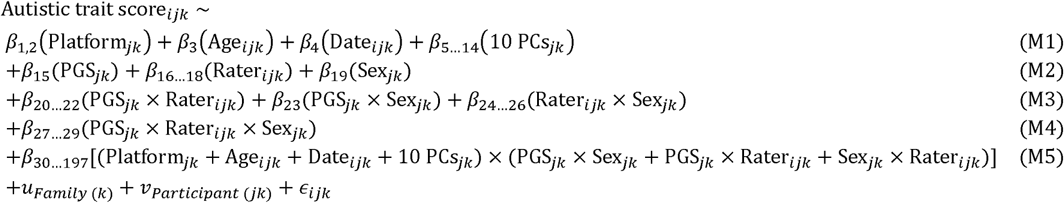

We compared hierarchical models using the Akaike information criterion (AIC), and likelihood-ratio tests against *p* = .0125 (Bonferroni-corrected for four independent tests).

### Post hoc analyses

We chose the best performing model for post hoc investigations. We further investigated specific effects within the two- and three-way interactions (such as comparisons between two specific raters) via estimated marginal means, using the *emmeans* R package^45^. For a full explanation, see Supplementary text.

Next, we calculated pairwise contrasts between the sexes within each rater, and between the raters within the sexes, in odds ratios.

Finally, we derived predicted probabilities of belonging to each specific autistic trait score level (*no, low, high*) for higher resolution interpretability and visualisation of results.

We controlled for multiple testing using false discovery rate (FDR) corrections within each post hoc analysis (i.e. each set of marginal means, pairwise contrasts between the sexes or raters, and predicted probabilities). We tested for significance at *α* = .05.

### Measurement invariance

Comparing scores between males and females is meaningful only if the items measure this latent trait in the same way in both sexes, a property known as measurement invariance (MI)^46^. MI holds when males and females with the same level of the latent trait have the same expected score on each item. We performed MI with respect to sex on the items of the So et al.^31^ autism scale per rater, as outlined in van der Sluis et al.^47^ and Millsap and Yun-Tein^48^. MI was tested by fitting a sequence of nested confirmatory factor analysis (CFA) models as visualised in Figure 1, and laid out in Table S1. See Supplementary text and Figure S2 for full description.

**Figure 1:**
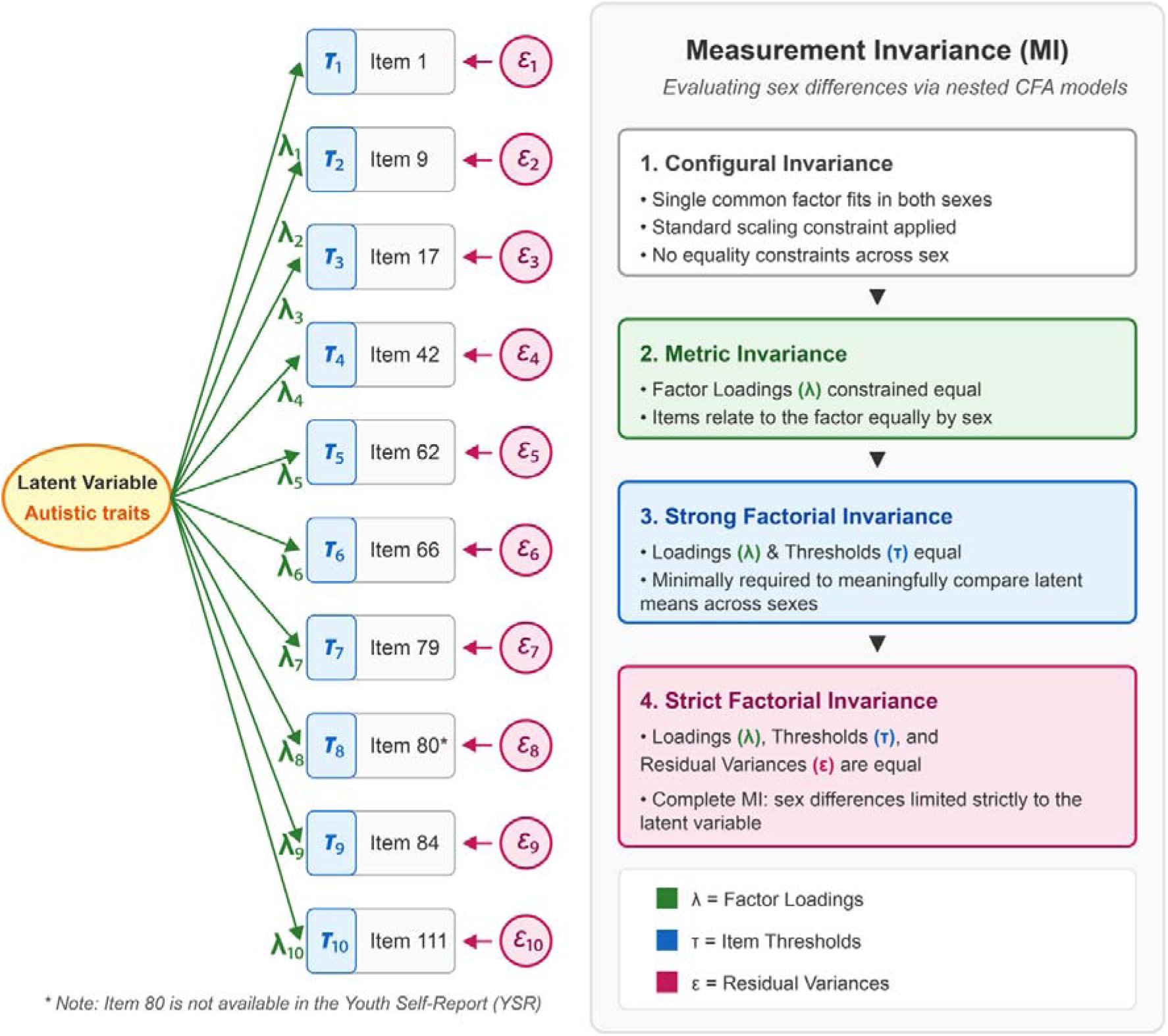
Measurement invariance. Two-group confirmatory factor analysis (CFA) models were fitted across sex, with a latent factor for autistic traits loading on the items of the So et al. (31) Achenbach System of Empirically Based Assessment (ASEBA) autistic traits subscale. Nested models with increasing equality constraints across sex on loadings and thresholds were compared to test measurement invariance. Not shown is the final model, in which the latent factor means were constrained to be equal across sexes to test for a sex difference in the latent trait.

## Results

### Sample descriptives

Data from up to four raters were available for 6,163 participants. **Table 1** shows descriptives by rater and sex, and **Figure 2** shows score distributions. Trait score correlations between the raters were low to moderate (.21–.64), highest between parents, and lowest between external and self-ratings, especially teacher and self-ratings (Table S2).

**Figure 2:**
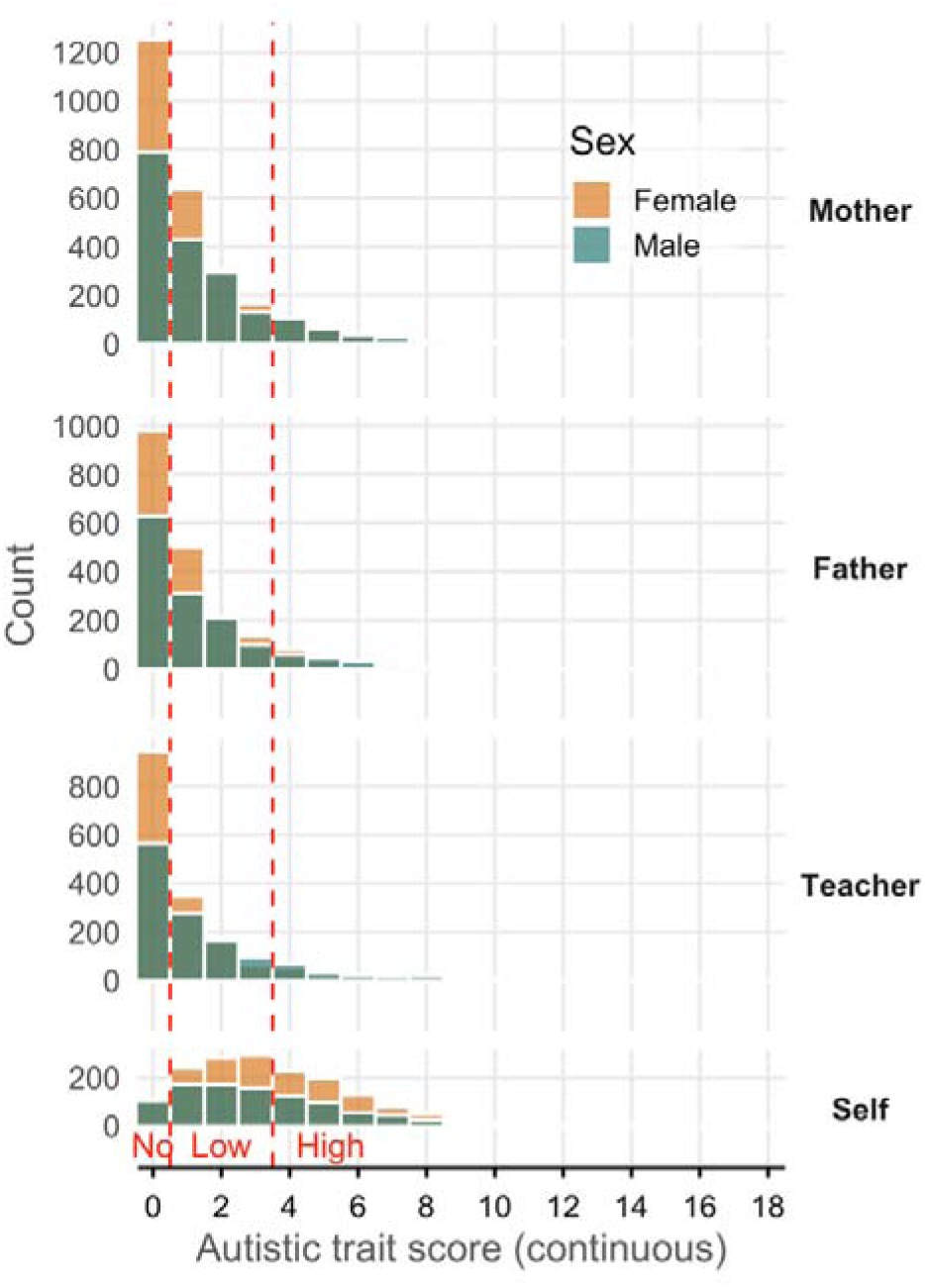
Histogram of autistic trait scores separated on rater and sex. Possible scores range 0–20. Red lines indicate cut-offs for ordinal categories *no*, *low*, and *high* at scores of 0, 1– 3, 4+. Male and female bins are overlapping, not stacked.

**Table 1:** Sample descriptives.

| **Rater** | **Sex** | **Sample size** | **Effective sample size** | **Age mean (*SD*)** | **Autistic trait score mean (*SD*)** |
|---|---|---|---|---|---|
| Mother | Male | 1,919 | 1,454 | 12.22 (0.36) | 1.55 (2.07) |
| Mother | Female | 2,622 | 1,940 | 12.21 (0.38) | 1.19 (1.70) |
| Father | Male | 1,420 | 1,045 | 12.22 (0.37) | 1.41 (1.93) |
| Father | Female | 1,991 | 1,491 | 12.19 (0.37) | 1.10 (1.64) |
| Teacher | Male | 1,292 | 956 | 12.13 (0.28) | 1.60 (2.30) |
| Teacher | Female | 1,647 | 1,293 | 12.13 (0.30) | 0.96 (1.72) |
| Self | Male | 979 | 826 | 14.80 (0.67) | 3.18 (2.41) |
| Self | Female | 1,653 | 1,288 | 14.76 (0.71) | 3.52 (2.37) |

Note: Autistic trait score is on continuous scale. Participants partly overlap between the different raters. Because clustering reduces the amount of independent information, we calculated effective sample sizes per sex and rater from intraclass correlation coefficients<sup>49</sup> (see GitHub for calculation). Abbreviations: *SD*: standard deviation.

For full sample and PGS descriptives, see Tables S3–S4. Histograms of age and assessment date separated by rater and sex are shown in Figures S3–S4.

### Ordinal regression model results

For full model estimates, see Tables S5–S11. Here, PGS and covariate effects are reported from model 3 after FDR correction (Table S7). Autism PGS were significantly associated with a higher autistic trait score level (*OR* = 1.22, 95% CI [1.15, 1.29], *p* < .001; i.e. 22% higher odds of belonging to a higher score level per 1 *SD* increase in the PGS). Age showed no significant effect (*OR* = 1.05, 95% CI [0.95, 1.16], *p* = .351), and assessment date showed a small but significant effect (*OR* = 1.01, 95% CI [1.00, 1.03], *p* = .033) such that later date resulted in higher score level.

In a linear regression with one participant per family, mean PGS did not differ between the sexes ( = 0.00, *SE* = 0.04, *p* = .943, *n* = 3226).

Hierarchical model comparisons (Table 2) identified model 3 as the best-fitting model according to the AIC and the likelihood-ratio test. The rater × sex interaction improved fit most (*χ*^2^(3) = 88.10, *p* < .001). In contrast, PGS × rater (*χ*^2^(3) = 1.97, *p* = .578) and PGS × sex (*χ*^2^(1) = 0.15, *p* = .700) did not significantly influence fit. The addition of the three-way interaction in model 4 worsened the AIC and did not significantly improve the likelihood-ratio (*p* = .845). Model 5 improved the likelihood-ratio (*p* < .001), but not the AIC (201 vs 33 parameters). Following these results, we report model 3 for main effects and two-way interactions, model 4 for the three-way interaction, and model 5 as a sensitivity analysis.

**Table 2:** Ordinal mixed model fits.

| **Model** | **n par. (*df*)** | **AIC** | **LogLik** | **LR χ<sup>2</sup> (*p*-value)** | ***R²* (marg)** | ***R²* (cond)** |
|---|---|---|---|---|---|---|
| 1: Base model | 18 | 24598 | -12,281 | — | .154 | .476 |
| 2: + Main effects | 23 (5) | 24211 | -12,083 | 396.44 (<.001<sup>a</sup>) | .178 | .489 |
| 3: + Two-way interactions | 30 (7) | 24135 | -12,037 | 90.84 (<.001<sup>a</sup>) | .183 | .495 |
| 4: + Three-way interaction | 33 (3) | 24140 | -12,037 | 0.82 (.845) | .183 | .495 |
| 5: + Covariate interactions | 201 (168) | 24198 | -11,898 | 278.07 (<.001<sup>a</sup>) | .205 | .515 |

Note: Hierarchical model comparison results with each model tested against the previous. Base model contains random effects and covariates. The table includes Nakagawa *R*<sup>2</sup> (Supplementary text) adapted for mixed models, with marginal (marg) representing the variance explained by fixed effects and conditional (cond) by fixed + random effects. Abbreviations: number of parameters with degrees of freedom (n par. (*df*)); Akaike information criterion (AIC); log-likelihood (LogLik); likelihood ratio chi-squared (LR χ<sup>2</sup>). <sup>a</sup>Significant after Bonferroni correction.

### Raters differentially rate males and females in their autistic trait scores

Post hoc contrasts of the rater × sex interaction (FDR-corrected; Tables S12–S13) showed higher autistic trait score levels for males in mother- (*OR* = 1.56, 95% CI [1.34, 1.83], *p* < .001), father- (*OR* = 1.49, 95% CI [1.25, 1.77], *p* < .001) and teacher-ratings (*OR* = 2.34, 95% CI [1.94, 2.84], *p* < .001). In contrast, females rated themselves higher (*OR* = 0.71 (males as reference group), 95% CI [0.59, 0.87], *p* < .001).

When contrasting raters within each sex, the self-rated scores were significantly higher than external raters’ scores (Table S14). Additionally, teachers scored females lower than mother (*OR* = 1.69, 95% CI [1.46, 1.96], *p* < .001) and fathers (*OR* = 1.54, 95% CI [1.32, 1.80], *p* < .001), but rated males similarly to parents. Mothers and fathers rated both sexes similarly.

Predicted probabilities of each score level mirrored these contrasts (**Figure 3**, Tables S15–S17). For mother, father, and teacher raters, males more often had *high* score levels, with the largest difference in teachers. For self-rated, the opposite was true, such that females more often had *high* score level.

**Figure 3:**
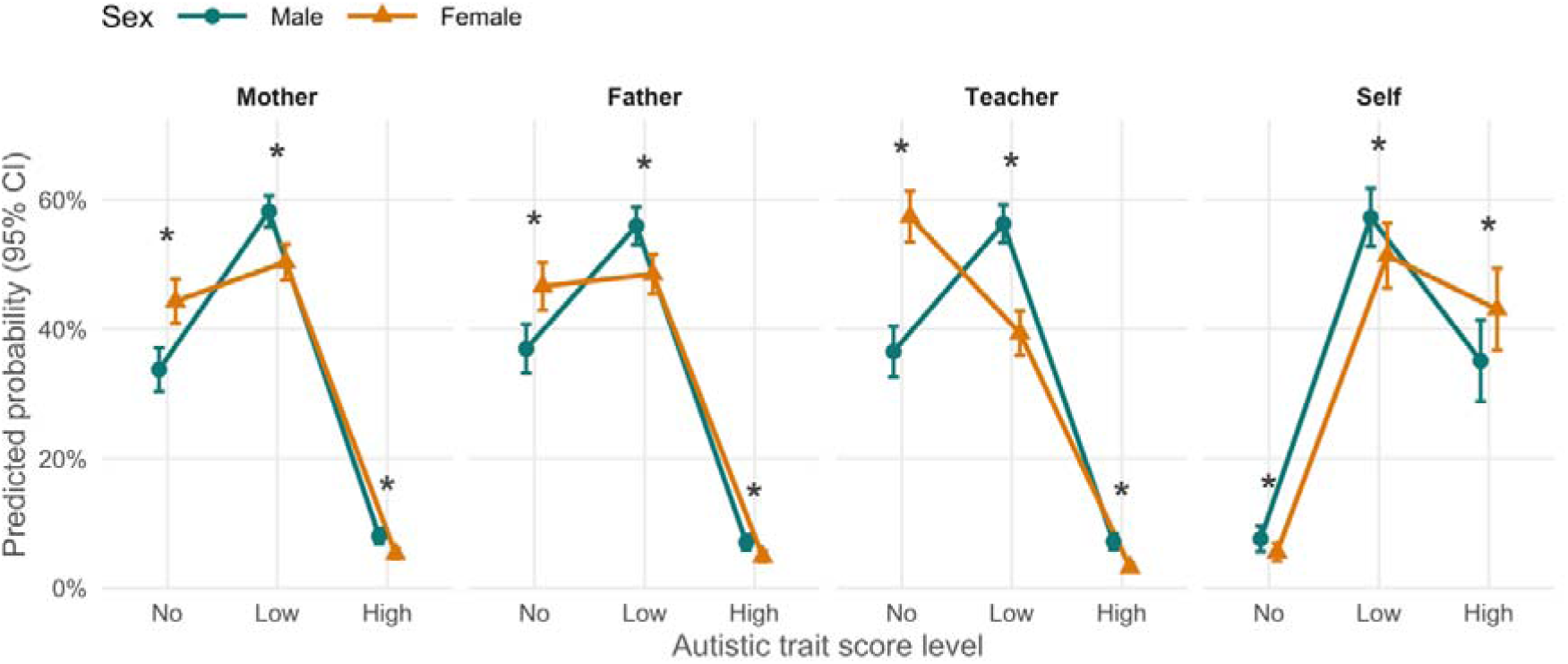
Autistic trait score by rater and sex. Predicted probability of belonging to an autistic trait score level. Probabilities were calculated at mean polygenic score (PGS) = 0. Data were extracted from model 3. All contrasts between males and females within a score level were significant at = .05 after false discovery rate (FDR) correction. Error bars are in 95% confidence intervals (CIs). Females as rated by mothers, fathers, and teachers are predicted to more often belong to the *no* score level, and less often to the *low* and *high* levels. In contrast, in self-ratings, females belong more often to the *high* score level. Indeed, on the latent scale, external raters significantly rated males higher, but females rated themselves higher.

### No sex differences in PGS association per rater

We estimated autism PGS associations with autistic trait score for each rater-sex combination (PGS × rater × sex; **Figure 4**, Tables S18–S20). The PGS was significantly associated in all rater-sex combinations (*ORs* = 1.17–1.31, *p* < .001– .020 after FDR correction). However, these associations did not differ between raters within either sex, or between the sexes within any rater (*ORs* = 0.91–1.09, *p* = .89–.93). These results indicate that the association between PGS and autistic trait score does not differ across sex, and different raters do not show different genetic signal.

**Figure 4:**
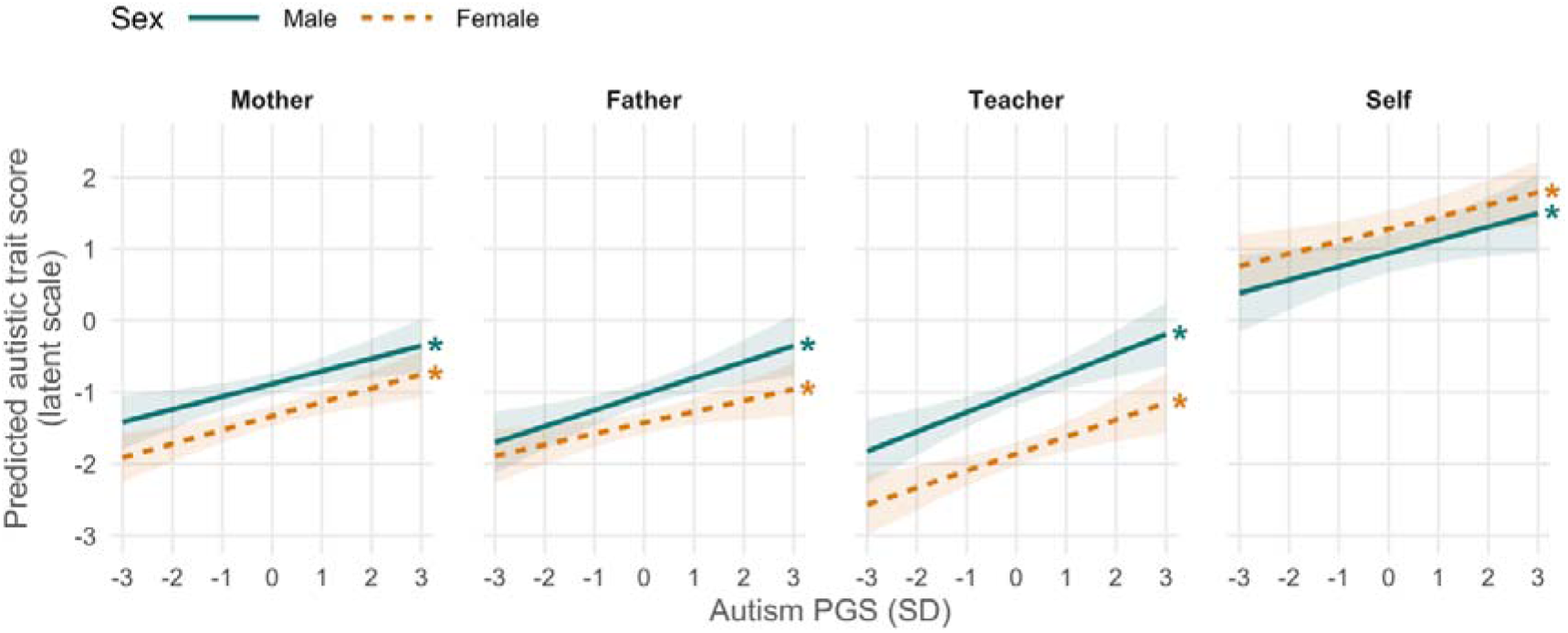
Autism PGS associations across rater and sex. Modeled by a three-way interaction of autism polygenic score (PGS) × rater × sex. Predicted autistic trait score on the latent scale of the ordinal model: an assumed continuous variable underlying the three score levels (*no*, *low*, *high*), in log-odds units. The origin of this scale is arbitrary, so only slopes and differences are interpretable. Based on model 4. Slopes include banded 95% confidence intervals (CIs). Asterisks at the end of a trend line indicate trends that differ from zero at false discovery rate (FDR)-corrected *p* < .05 (indicating for all raters a main effect of PGS).

### Measurement invariance shows a sex bias in item 1

In the MI analysis of the So et al.^31^ autism scale, item 1 – *“Acts too young for his/her age”* – showed a sex bias towards males in mother- and self-raters. As between the ages of 12–14 males and females show a differing development^50^, it is reasonable to assume that this is reflected in the item. For teacher and father raters, no substantial item biases were found. Detailed results can be found in Supplementary text and Tables S21–S22.

### Sensitivity analyses

We performed several sensitivity analyses.

First, removing item 80 (absent from the YSR) and item 1 (sex-biased according to MI) from the regression analyses did not change the results (Tables S23–S26). To rule out sample composition effects, we reran the models in participants with all four raters present (male *n* = 346, female *n* = 556; Tables S27–S30). The pattern of results was comparable, although the higher self-ratings in females no longer reached significance (*OR* = 0.81, 95% CI [0.58, 1.13], *p* = .214). Sex differences across raters persisted at a PGS 2 *SD* above the mean, reflecting participants at a high genetic liability as assumed under the LTM (Table S31).

In model 5 (adding covariate interactions), results were largely unchanged. However, the self-rated sex difference was no longer significant, although its point estimate was similar (*OR* = 0.69 vs 0.71; Supplementary text). Because self-ratings were collected at age 14 and external ratings at age 12, rater and age effects are confounded. In a YNTR subsample with self-reports at age 12 (male *n* = 337, female *n* = 382), we found no sex difference in self-rated autistic trait scores (*OR* = 1.03, 95% CI [0.69, 1.55], *p* = .881), although the wide CI reflects the small sample.

## Discussion

We investigated the interplay between autism PGS, rater, and sex on ASEBA-based autistic traits from four raters in a general population sample. Mothers, fathers, and teachers rated males higher than females, whereas females rated themselves higher, consistent with under-recognition of autistic traits in females by external raters. The autism PGS was associated with autistic traits, albeit with a small effect size^51^, but this association did not differ by sex, rater, or their combination, providing no evidence for sex amplification. One item showed a sex bias in mother- and self-ratings.

Our findings can be interpreted under the Liability Threshold Model (LTM), where autism is diagnosed when an underlying continuous liability, to which genetic and environmental factors contribute, exceeds a threshold^52^. The LTM is a possible framework to explain the diagnostic sex ratio^8,52^. A higher mean PGS in autistic females (without an intellectual disability) as we previously found^14^ implies that females need to carry more autism-associated common variants before crossing the threshold. There are two mechanisms that could produce this (for a more detailed description see Supplementary text and Figures S5–S6). First, under sex amplification, the same variants would have larger effects in males^15^, so that at the same PGS females show lower trait levels and the PGS association is weaker in females. Alternatively, variant effects are equal, but the threshold is higher for females^52^, for instance because autistic traits in females need to be more pronounced before they are recognised. Our results fit the second mechanism better. The PGS association did not differ by sex for any rater. In contrast, external raters reported markedly higher trait levels in males, also at a PGS 2 *SD* above the mean — the upper tail most relevant to the LTM. This is consistent with a scenario in which under-recognition by external raters means that only females with higher trait levels, and correspondingly a higher PGS, receive a diagnosis.

Importantly, current autism PGS capture only a limited proportion of the genetic effects on autism, and the majority of genetic effects are thus not represented in this study. The small effect size of the autism PGS in our analysis may limit the ability to detect subtle sex differences in genetic associations with trait expression. Although the common-variant association with autistic traits appeared similar across sexes and raters, rare and de novo variants, X-chromosomal effects, and non-additive effects could differ.

Clinical use of PGS is of growing interest yet remains controversial^53^. The effect size of the autism PGS in our results and in referenced research does not yet support clinical use in individual diagnosis; rather they remain a population-level research tool^11,54^. Our findings suggest that the rater choice is unlikely to matter for capturing the current autism PGS signal, informing future use. For genetic studies of autistic traits this means that the rater with the largest available sample can be used, provided that rater differences in mean trait levels between males and females are accounted for.

Discrepancies between internal and external raters in autism have repeatedly been reported^18,19,21,22^ and are often explained through differences in trait expression between the sexes. Autistic females often show more internalised autistic traits, behaviour, and co-occurring conditions^6,55,56^, whereas autistic males score higher on externalising traits and behaviours that are more noticeable to their immediate environment^57–59^. However, findings on behavioural differences are often inconsistent, and while subtle differences likely exist, the sexes largely overlap in their core traits^6,21,60^. Another explanation currently discussed is masking, where the autistic individual attempts to hide their autistic traits to appear more neurotypical for their immediate surroundings^61,62^. Females generally mask more than males^61^. Our results could be explained through both differences in autistic traits and masking behaviour of females. The under-recognition of autistic traits is associated with a later diagnosis^21,57^, and worse mental health outcomes for females^57,62^. This highlights the need for an increased understanding of autistic traits in females.

Teachers often play an important role in autism diagnosis^63,64^. In our results, we found a more extreme sex discrepancy in autistic trait scores in teacher ratings compared to mother and father ratings. This is in line with earlier work by Putnam et al.^19^, who found no sex differences in parent-reported autistic traits but reported males as having higher traits in teacher reports as measured by the Social Responsiveness Scale. In a vignette study by Whitlock et al.^64^, descriptions of autistic boys and girls were given to primary school teachers, who showed bias against the girl description. These findings indicate that teachers might especially under-recognise autistic traits in females, and implicate the school system as a societal barrier for females in reaching a diagnosis.

It is important to note that no single rater can serve as a gold standard^65^. Self-ratings have unique access to internalised traits, yet individuals may be less able to judge their own social behaviour than external raters are. Teachers can more easily compare their students against peers but may miss an internalised phenotype. Parents observe their children in more diverse and natural contexts yet have fewer peers to compare against. Investigating the sources of rater discrepancies is complex and of interest for future studies.

## Limitations

Several limitations should be considered when interpreting our results. First, the autism subscale was validated by So et al.^31^ for the CBCL and TRF, but not for the YSR, which lacks one of the CBCL items. Removing the item from the other scales did not change our results.

Second, participants were assessed in narrow age ranges: around 12 for external raters, and 14 for self-reports. As autism is a neurodevelopmental condition, it would be valuable to compare autistic traits across different ages. Because self-reports were collected at a different age than the other ratings, we could not categorically distinguish rater effects from age effects. How age differentially influences the way male and female adolescents rate their own autistic traits remains an open question.

Finally, our sample was restricted to European ancestry. This limits how results generalise, and we are likely to have missed how autistic traits are viewed and experienced in other populations or cultures.

## Conclusion

Using the same autism subscale across four raters, we found that mothers, fathers, and teachers rated males higher than females, whereas females rated themselves higher. However, this difference could be confounded with age. The autism PGS association did not differ by sex or rater, providing no evidence for sex amplification, although modest amplification cannot be excluded at current PGS effect sizes. As the associations did not differ between raters, genetic studies of autistic traits may pragmatically choose the rater with the largest available sample. External raters, particularly teachers, may overlook autistic traits in females, possibly contributing to the diagnostic sex bias.

## Supplementary files

### GitHub

https://github.com/technologiemas/PGS_association_study_autism_score

## Declarations

### Author contributions: CRediT

**TS** contributed to Conceptualization, Data Curation, Formal Analysis, Investigation, Methodology, Project Administration, Software, Visualization, Writing – original draft. **MMW** contributed to Writing – review & editing. **RP** contributed to Data Curation, Investigation, Software, Writing – review & editing. **MB** contributed to Data Curation, Investigation, Resources. **TJCP** contributed to Conceptualization, Methodology, Supervision, Writing – review & editing, Funding acquisition. **WAA** contributed to Conceptualization, Methodology, Supervision, Writing – review & editing.

## Data Availability

Summary data produced in the analyses are contained in the manuscript. Raw individual data is available upon request to the Netherlands Twin Register.

https://ntrdata.nl/data-showcase.html

## Acknowledgements

We kindly thank the Netherlands Twin Register for data access, and the YNTR participants for contributing their data. We wish to express special thanks to Conor V. Dolan for his help with statistical modelling and measurement invariance.

Autism Working Group of the Psychiatric Genomics Consortium: Richard J.L. Anney^4^, Anders Børglum^5,6,7^, Angel Carracedo^8,9,10,11^, Elizabeth C. Corfield^12,13,14,15^, Jinjie Duan^5,6,7^, Isabel Dziobek^16^, Javier González-Peñas^17,18^, Magdalena Janecka^19^, Susan S. Kuo^20,21,22^, Stephan J. Sanders^23,24^, Stephen W. Scherer^25,26^, Ludger Tebartz van Elst^27^, Mohammed Uddin^28,29^, Jack F.G. Underwood^4^, Jacob A.S. Vorstman^30,31,32^, Varun Warrier^33^, Lauren A. Weiss^34^.

^4^Department of Psychological Medicine and Clinical Neurosciences, Cardiff University, Cardiff, UK

^5^Department of Biomedicine, Aarhus University, Aarhus, Denmark

^6^Lundbeck Foundation Initiative for Integrative Psychiatric Research (iPSYCH), Aarhus, Denmark

^7^Center for Genomics and Personalized Medicine, Aarhus, Denmark

^8^Grupo de Medicina Xenómica (CiMUS), Universidade de Santiago de Compostela (USC), Santiago de Compostela, Spain

^9^Fundación Pública Galega de Medicina Xenómica (FPGMX), Servizo Galego de Saúde (SERGAS), Santiago de Compostela, Spain

^10^Grupo de Genética, Instituto de Investigación Sanitaria de Santiago de Compostela (IDIS), Santiago de Compostela, Spain

^11^Centro de Investigación Biomédica en Red de Enfermedades Raras (CIBERER), Instituto de Salud Carlos III, Madrid, Spain

^12^PsychGen Centre for Genetic Epidemiology and Mental Health, Norwegian Institute of Public Health, Oslo, Norway

^13^Psychiatric Genetic Epidemiology Group, Research Department, Lovisenberg Diaconal Hospital, Oslo, Norway

^14^MRC Integrative Epidemiology Unit, Bristol Medical School, University of Bristol, Bristol, UK

^15^Population Health Sciences, Bristol Medical School, University of Bristol, Bristol, UK

^16^Department of Psychology, Humboldt-Universität zu Berlin, Germany

^17^Department of Child and Adolescent Psychiatry, Institute of Psychiatry and Mental Health, Hospital General Universitario Gregorio Marañón, Instituto de Investigación Sanitaria Gregorio Marañón (IiSGM), Madrid, Spain

^18^CIBER de Salud Mental, Instituto de Salud Carlos III, Madrid, Spain

^19^Department of Child and Adolescent Psychiatry, NYU Langone, New York, NY, USA

^20^Department of Psychiatry, Massachusetts General Hospital, Harvard Medical School, Boston, MA, USA

^21^Center for Genomic Medicine, Massachusetts General Hospital, Boston, MA, USA

^22^Stanley Center for Psychiatric Research, Broad Institute of MIT and Harvard, Cambridge, MA, USA

^23^Institute of Developmental and Regenerative Medicine, Department of Paediatrics, University of Oxford, Oxford, UK

^24^Department of Psychiatry and Behavioral Sciences, UCSF Weill Institute for Neurosciences, University of California San Francisco, San Francisco, CA, USA

^25^The Centre for Applied Genomics and Genetics and Genomic Biology Program, The Hospital for Sick Children, Toronto, ON, Canada

^26^McLaughlin Centre and Department of Molecular Genetics, University of Toronto, Toronto, ON, Canada

^27^Department of Psychiatry and Psychotherapy, Medical Center – University of Freiburg, Faculty of Medicine, Freiburg, Germany

^28^Center for Applied and Translational Genomics (CATG), Mohammed Bin Rashid University of Medicine and Health Sciences (MBRU), Dubai Health, Dubai, UAE

^29^GenomeArc Inc., Mississauga, ON, Canada

^30^Department of Psychiatry, The Hospital for Sick Children, Toronto, ON, Canada

^31^Department of Psychiatry, Temerty Faculty of Medicine, University of Toronto, Toronto, ON, Canada

^32^Program in Genetics and Genome Biology, SickKids Research Institute, The Hospital for Sick Children, Toronto, ON, Canada

^33^Department of Psychiatry, University of Cambridge, Cambridge, UK

^34^Institute for Human Genetics and Department of Psychiatry and Behavioral Sciences, UCSF Weill Institute for Neurosciences, University of California San Francisco, San Francisco, CA, USA

## Statements and declarations

Anders Børglum reports having received a speaker’s fee from Medice Nordic. Stephan J. Sanders receives research funding from BioMarin Pharmaceutical. Stephen W. Scherer served on the Scientific Advisory Committee of Population Bio, Deep Genomics and Diploid Genomics; intellectual property from aspects of his research held at The Hospital for Sick Children is licensed to Athena Diagnostics and Population Bio. Jacob A.S. Vorstman serves as a consultant for Synaptiq Therapeutics for the design of a clinical trial in children with the 22q11.2 deletion. Varun Warrier receives research funding from Centile Biosciences. All other authors report no conflicts of interest.

Large language models (LLMs) were used in this project. Microsoft Copilot and Anthropic’s Claude Code assisted with code completion, debugging and limited code generation; generated code is marked with comments. Google Gemini and Claude were used for brainstorming and as conceptual support for narrative structure and statistical modelling. Claude was also used to check formatting against APA and journal guidelines, and to check consistency of terminology and spelling. Beyond occasional suggestions of synonyms or rewrites, LLMs were not used to draft manuscript text. The authors reviewed all AI-assisted output and take full responsibility for the content.

## Funding and registration

This study was conducted as part of the NWA-ORC-SCANNER consortium, funded by the Dutch Research Council (NWO), grant: NWA.1518.22.136; DOI: https://doi.org/10.61686/PSQSK66981. TJCP is supported by ZonMW under grant number 60-63600-98-834. WAA is supported by a Dutch Ministry of Education, Culture and Sciences (OCW) Talent and Early Development Grant. Preregistration for this manuscript was done via Open Science Framework (OSF) under DOI: 10.17605/OSF.IO/4PQEB.

## Notes

### Author Declarations

The Netherlands Twin Register gave ethical approval for this research. Data collection received approval from the Central Ethics Committee on Research Involving Human Subjects at the Amsterdam University Medical Centres.

## References

1. Lord, C. et al. Autism spectrum disorder. Nat. Rev. Dis. Primers 6, 5 (2020).

2. Loomes, R., Hull, L. & Mandy, W. P. L. What Is the Male-to-Female Ratio in Autism Spectrum Disorder? A Systematic Review and Meta-Analysis. J. Am. Acad. Child Adolesc. Psychiatry 56, 466–474 (2017).

3. Zeidan, J. et al. Global prevalence of autism: A systematic review update. Autism Res. 15, 778–790 (2022).

4. Ferri, S. L., Abel, T. & Brodkin, E. S. Sex Differences in Autism Spectrum Disorder: a Review. Curr. Psychiatry Rep. 20, 9 (2018).

5. Lockwood Estrin, G., Milner, V., Spain, D., Happé, F. & Colvert, E. Barriers to Autism Spectrum Disorder Diagnosis for Young Women and Girls: a Systematic Review. *Rev*. J. Autism Dev. Disord. 8, 454–470 (2021).

6. Lai, M.-C., Lombardo, M. V., Auyeung, B., Chakrabarti, B. & Baron-Cohen, S. Sex/Gender Differences and Autism: Setting the Scene for Future Research. J. Am. Acad. Child Adolesc. Psychiatry 54, 11–24 (2015).

7. Halladay, A. et al. Sex and gender differences in autism spectrum disorder: summarizing evidence gaps and identifying emerging areas of priority. Mol. Autism 6, 36 (2015).

8. Sandin, S. et al. Examining Sex Differences in Autism Heritability. JAMA Psychiatry 81, 673–680 (2024).

9. Tick, B., Bolton, P., Happé, F., Rutter, M. & Rijsdijk, F. Heritability of autism spectrum disorders: a meta-analysis of twin studies. J. Child Psychol. Psychiatry 57, 585–595 (2016).

10. Kullo, I. J. et al. Polygenic scores in biomedical research. Nat. Rev. Genet. 23, 524–532 (2022).

11. Furrer, R. A. & Turkheimer, E. N. Polygenic risk scores are not genetic predispositions. *Nat*. Hum. Behav. 10, 1177–1178 (2026).

12. Askeland, R. B. et al. Early manifestations of genetic risk for neurodevelopmental disorders. J. Child Psychol. Psychiatry 63, 810–819 (2022).

13. Serdarevic, F. et al. Polygenic risk scores for developmental disorders, neuromotor functioning during infancy, and autistic traits in childhood. Biol. Psychiatry 87, 132–138 (2020).

14. Sollie, T., Akingbuwa, W. A., de Wit, M. M., Badura, A. & Polderman, T. J. C. Sex Differences in the role of Additive Genetic Variants in Autism: A Systematic Review. medRxiv 2025.12.23.25342905 (2025) doi:10.64898/2025.12.23.25342905.

15. Zhu, C. et al. Amplification is the primary mode of gene-by-sex interaction in complex human traits. Cell Genom. 3, 100297 (2023).

16. Crane, L., Chester, J. W., Goddard, L., Henry, L. A. & Hill, E. Experiences of autism diagnosis: A survey of over 1000 parents in the United Kingdom. Autism 20, 153–162 (2016).

17. Hosozawa, M. et al. Determinants of an autism spectrum disorder diagnosis in childhood and adolescence: Evidence from the UK Millennium Cohort Study. Autism 24, 1557–1565 (2020).

18. De la Marche, W. et al. Measuring quantitative autism traits in families: informant effect or intergenerational transmission? Eur. Child Adolesc. Psychiatry 24, 385– 395 (2015).

19. Putnam, O. C., McFayden, T. C. & Harrop, C. Sex Differences and Parent– Teacher Discrepancies in Reports of Autism Traits: Evidence for Camouflaging in a School Setting. J. Autism Dev. Disord. 55, 3888–3899 (2025).

20. Ronald, A., Happé, F. & Plomin, R. A twin study investigating the genetic and environmental aetiologies of parent, teacher and child ratings of autistic-like traits and their overlap. Eur. Child Adolesc. Psychiatry 17, 473–483 (2008).

21. Cruz, S. et al. Is There a Bias Towards Males in the Diagnosis of Autism? A Systematic Review and Meta-Analysis. Neuropsychol. Rev. 35, 153–176 (2025).

22. Terner, M. & Golan, O. Sex Differences in Adult Autism Screening: A Comparison of Current Self-Report and Retrospective Parent-Report Measures. J. Autism Dev. Disord. 56, 3205–3214 (2025).

23. Riglin, L. et al. Investigating attention-deficit hyperactivity disorder and autism spectrum disorder traits in the general population: What happens in adult life? J. Child Psychol. Psychiatry 62, 449–457 (2021).

24. Bottema-Beutel, K., Kapp, S. K., Lester, J. N., Sasson, N. J. & Hand, B. N. Avoiding Ableist Language: Suggestions for Autism Researchers. Autism Adulthood 3, 18–29 (2021).

25. Buijsman, R., Begeer, S. & Scheeren, A. M. ‘Autistic person’ or ‘person with autism’? Person-first language preference in Dutch adults with autism and parents. Autism 27, 788–795 (2023).

26. R Core Team. R: A Language and Environment for Statistical Computing. https://www.R-project.org/ (2025).

27. Muthén, L. K. & Muthén, B. Mplus User’s Guide. (Los Angeles, CA: Muthén & Muthén, 2017).

28. Ligthart, L. et al. The Netherlands Twin Register: Longitudinal Research Based on Twin and Twin-Family Designs. Twin Res. Hum. Genet. 22, 623–636 (2019).

29. van Beijsterveldt, C. E. M. et al. The Young Netherlands Twin Register (YNTR): Longitudinal Twin and Family Studies in Over 70,000 Children. Twin Res. Hum. Genet. 16, 252–267 (2013).

30. Achenbach, T. M. & Rescorla, L. A. Manual for the ASEBA School-Age Forms & Profiles. (University of Vermont, Research Center for Children, Youth, & Families, Burlington, VT, 2001).

31. So, P. et al. Using the Child Behavior Checklist and the Teacher’s Report Form for identification of children with autism spectrum disorders. Autism 17, 595–607 (2013).

32. Grove, J. & Zhang, J. Advancing our understanding of the genetics of autism: findings from the largest GWAS to date. Eur. Neuropsychopharmacol. 111, 112956 (2026).

33. Zeng, J. et al. Signatures of negative selection in the genetic architecture of human complex traits. Nat. Genet. 50, 746–753 (2018).

34. Zheng, Z. et al. Leveraging functional genomic annotations and genome coverage to improve polygenic prediction of complex traits within and between ancestries. Nat. Genet. 56, 767–777 (2024).

35. Chang, C. C. et al. Second-generation PLINK: rising to the challenge of larger and richer datasets. GigaScience 4, s13742-015-0047–8 (2015).

36. Purcell, S. & Chang, C. PLINK2.

37. Mansolf, M., Blackwell, C. K., Cummings, P., Choi, S. & Cella, D. Linking the Child Behavior Checklist to the Strengths and Difficulties Questionnaire. Psychol. Assess. 34, 233–246 (2022).

38. Kuzminski, R. et al. Linking knowledge and attitudes: Determining neurotypical knowledge about and attitudes towards autism. PLoS ONE 14, e0220197 (2019).

39. Fyfe, C. et al. Time trends in the male to female ratio for autism incidence: population based, prospectively collected, birth cohort study. BMJ 392, e084164 (2026).

40. Price, A. L. et al. Principal components analysis corrects for stratification in genome-wide association studies. Nat. Genet. 38, 904–909 (2006).

41. Choi, S. W., Mak, T. S.-H. & O’Reilly, P. F. Tutorial: a guide to performing polygenic risk score analyses. Nat Protoc 15, 2759–2772 (2020).

42. Hoaglin, D. C., Iglewicz, B. & Tukey, J. W. Performance of Some Resistant Rules for Outlier Labeling. J. Am. Stat. Assoc. 81, 991–999 (1986).

43. Christensen, R. H. B. ordinal—regression models for ordinal data. (2023).

44. Keller, M. C. Gene-by-environment interaction studies have not properly controlled for potential confounders: The problem and the (simple) solution. Biol. Psychiatry 75, 18–24 (2014).

45. Lenth, R. V. Emmeans: Estimated Marginal Means, Aka Least-Squares Means. https://rvlenth.github.io/emmeans/ (2025).

46. Putnick, D. L. & Bornstein, M. H. Measurement invariance conventions and reporting: The state of the art and future directions for psychological research. Dev. Rev. 41, 71–90 (2016).

47. van der Sluis, S. et al. Sex differences and gender-invariance of mother-reported childhood problem behavior. Int. J. Methods Psychiatr. Res. 26, e1498 (2017).

48. Millsap, R. E. & Yun-Tein, J. Assessing Factorial Invariance in Ordered-Categorical Measures. Multivariate Behav. Res. 39, 479–515 (2004).

49. Guittet, L., Ravaud, P. & Giraudeau, B. Planning a cluster randomized trial with unequal cluster sizes: practical issues involving continuous outcomes. BMC Med Res Methodol 6, 17 (2006).

50. Marceau, K., Ram, N., Houts, R. M., Grimm, K. J. & Susman, E. J. Individual Differences in Boys’ and Girls’ Timing and Tempo of Puberty: Modeling Development With Nonlinear Growth Models. Dev. Psychol. 47, 1389–1409 (2011).

51. Chen, H., Cohen, P. & Chen, S. How Big is a Big Odds Ratio? Interpreting the Magnitudes of Odds Ratios in Epidemiological Studies. Commun. Stat. Simul. Comput. 39, 860–864 (2010).

52. Dougherty, J. D. et al. Can the “female protective effect” liability threshold model explain sex differences in autism spectrum disorder? Neuron 110, 3243–3262 (2022).

53. Kullo, I. J. Clinical use of polygenic risk scores: current status, barriers and future directions. Nat. Rev. Genet. 27, 246–263 (2026).

54. Straub, V. J. et al. Early-career-researcher-led best practices for social science and behavioural genetics. *Nat*. Hum. Behav. 10, 1014–1018 (2026).

55. Solomon, M., Miller, M., Taylor, S. L., Hinshaw, S. P. & Carter, C. S. Autism Symptoms and Internalizing Psychopathology in Girls and Boys with Autism Spectrum Disorders. J. Autism Dev. Disord. 42, 48–59 (2012).

56. Cardon, G., McQuarrie, M., Calton, S. & Gabrielsen, T. P. Similar overall expression, but different profiles, of autistic traits, sensory processing, and mental health between young adult males and females. Res. Autism Spectr. Disord. 109, 102263 (2023).

57. Lai, M.-C. et al. A Behavioral Comparison of Male and Female Adults with High Functioning Autism Spectrum Conditions. PLoS ONE 6, e20835 (2011).

58. Wood-Downie, H., Wong, B., Kovshoff, H., Cortese, S. & Hadwin, J. A. Research Review: A systematic review and meta-analysis of sex/gender differences in social interaction and communication in autistic and nonautistic children and adolescents. J. Child Psychol. Psychiatry 62, 922–936 (2021).

59. Edwards, H., Wright, S., Sargeant, C., Cortese, S. & Wood-Downie, H. Research Review: A systematic review and meta-analysis of sex differences in narrow constructs of restricted and repetitive behaviours and interests in autistic children, adolescents, and adults. J. Child Psychol. Psychiatry 65, 4–17 (2024).

60. Rubenstein, E., Wiggins, L. D. & Lee, L.-C. A Review of the Differences in Developmental, Psychiatric, and Medical Endophenotypes Between Males and Females with Autism Spectrum Disorder. J. Dev. Phys. Disabil. 27, 119–139 (2015).

61. Cook, J., Hull, L., Crane, L. & Mandy, W. Camouflaging in autism: A systematic review. Clin. Psychol. Rev. 89, 102080 (2021).

62. Scheeren, A. M., Nieuwenhuis, S., Crane, L., Roke, Y. & Begeer, S. Masking, social context and perceived stress in autistic adults: An ecological momentary assessment study. Autism 29, 3002–3013 (2025).

63. Conte, L. et al. Autism Spectrum Disorders and inclusion attitudes in the Italian school environments: teachers’ knowledge, attitudes, perceptions and their necessity to consult a healthcare multidisciplinary team. Acta Biomed. 93, e2022284 (2022).

64. Whitlock, A., Fulton, K., Lai, M.-C., Pellicano, E. & Mandy, W. Recognition of Girls on the Autism Spectrum by Primary School Educators: An Experimental Study. Autism Res. 13, 1358–1372 (2020).

65. De Los Reyes, A. & Epkins, C. C. Introduction to the Special Issue. A Dozen Years of Demonstrating That Informant Discrepancies are More Than Measurement Error: Toward Guidelines for Integrating Data from Multi-Informant Assessments of Youth Mental Health. J. Clin. Child Adolesc. Psychol. 52, 1–18 (2023).

