## Supplementary text for "The Interplay Between Sex, Rater, and Autism Polygenic Scores in Autistic Traits in General Population Adolescents"

### Deviations from preregistration

A preregistration for this study was published online on Open Science Framework ([DOI: 10.17605/OSF.IO/4PQEB](https://doi.org/10.17605/OSF.IO/4PQEB)). In the main manuscript, we diverge at several points from this preregistration, summarised below.

First, we preregistered two regression models, one with and one without interaction effects. However, to get more granular information about our results we decided to run a set of five hierarchical models so that main and interaction effects were introduced in a stepwise manner. For a comparison of the models, we decided to additionally include the Akaike information criterion as it considers model complexity. Second, we did not preregister the inclusion of assessment date as a covariate but decided to do so after a recent publication on period effects^1^. Finally, several sensitivity analyses were performed to investigate questions arising from our main results, which were not originally preregistered.

### Supplementary methods

#### Ethics

All research procedures complied with the ethical guidelines set by relevant institutional and national research committees, as well as the principles outlined in the Declaration of Helsinki. Data collection received approval from the Central Ethics Committee on Research Involving Human Subjects at the Amsterdam University Medical Centres. Informed consent was obtained from parents for all participants, including consent to approach of the teachers of the participants.

#### So et al. ASEBA autism subscale items

The So et al.^2^ ASEBA subscale consists of the following ten items for the CBCL and TRF. The YSR contains the same items but is missing item 80.

Item 1: *"Acts too young for his/her age"*

Item 9: *"Cannot get his/her mind off certain thoughts; obsessions"*

Item 17: *"Daydreams or gets lost in his/her thoughts"*

Item 42: *"Would rather be alone than with others"*

Item 62: *"Poorly coordinated or clumsy"*

Item 66: *"Repeats certain acts over and over"*

Item 79: *"Speech problem"*

Item 80: *"Stares blankly"**

Item 84: *"Strange behaviour"*

Item 111: *"Withdrawn, does not get involved with others"*.

#### Genotyping, quality control and PGS calculation

Participants were genotyped as part of the Netherlands Twin Register (NTR) study (version 18 of the NTR genotyping data), using three SNP arrays: Affymetrix 6.0 (N=1698), Affymetrix Axiom (N=494), and Illumina GSA NTR array (N=3971). Calling followed manufacturer protocols. Samples were checked for sex mismatches, heterozygosity outliers, and IBD-based pedigree mismatches, requiring ≥90% call rate overall and ≥80% per chromosome. SNP-level quality control (QC, applied per platform) required call rate >95%, HWE p>0.0001, MAF>0.01, and Mendelian/genotyping error rate <1%. After strand alignment and removal of ambiguous (palindromic) SNPs with MAF>0.30, each platform was imputed separately with *Beagle 5.4*^3^ against 1000 Genomes Phase 3^4^. Platform-imputed datasets were merged with *bcftools*^5^ into a single genotype data set for analysis. Genotype data were restricted to individuals with relevant, non-missing phenotype and covariate data prior to analysis.

We ran principal component analysis (PCA) for genetic ancestry estimation. Using QC'd SNPs common to 1000 Genomes Phase 3-imputed data across all three platforms (MAF>0.05, HWE p>0.001, call rate>0.98, Mendelian error <1%, imputation INFO≥0.90), SNPs were LD-pruned (*PLINK 1.9*, --indep 50 5 2)^6,7^ and long-range LD regions removed^8^, leaving 110,558 SNPs. PCs were computed in the 1000 Genomes reference panel and projected onto NTR samples with *smartpca*^9^ to identify and exclude ancestry outliers based on 1000 Genomes European reference individual values on principal component 1 up to 10.

For calculation of PGS we took the following steps: Discovery GWAS variant EAF and effect sizes were aligned with the NTR reference for the 1000 genomes variants. We retained variants for which the effect allele frequency (EAF) was 0.01 ≤ EAF ≤ 0.99. Discovery variants that were not part of this reference were discarded.

The *SBayesRC* method is implemented in the *gctb* software^10,11^. The method enables polygenic prediction by accounting for functional genomic annotations of over 7 million variants. By assuming the LD structure in the discovery GWAS is the same as in the (provided) reference, *SBayesRC* uses a low-rank approximation based on eigenvalues and eigenvectors of LD correlation matrices. The reference LD structure provided by the *SBayesRC* team is determined from a sample of N = 20,000 individuals from the UK Biobank (UKB). By selecting GWASs that include the European population, we adhere to this assumption. Moreover, the genetic similarity between UKB and NTR (the target population) ensure accurate prediction.

Since the LD structure in reference and discovery is assumed the same, summary statistics of missing variants in the discovery GWAS are imputed in a first step. The second step of the method involves actually calculating the weighted effect sizes.

#### Data cleaning

We excluded autistic trait score ratings with outlying ages using Tukey's fences^12^, i.e. ages more than 1.5 × the interquartile range below the lower quartile or above the upper quartile. To limit the age difference between raters, we only applied this one-sided: for mother-, father-, and teacher-ratings at age 12, we excluded ages below the lower fence (*n* = 0, 0, and 106, respectively), and for self-ratings at age 14, ages above the upper fence (*n* = 121).

#### Simulation to determine regression modelling

To determine which type of regression model to use we performed simulations. The autistic trait score data showed a non-normal distribution with a zero-inflated skew. We simulated this data using NTR scripts (available on [GitHub](https://github.com/technologiemas/PGS_association_study_autism_score)) and ran a mixed effect linear regression of the PGS predicting autistic trait score, with a random effect for family id. We used the *check_model, check_normality* and *check_heteroscedasticity* functions from the *performance* package^13^ to investigate posterior predictive check, homogeneity of variance and the normality of residuals (**Figure S1**), which were all violated. Additionally, the residuals showed high skew and kurtosis.

We concluded that linear regression violates multiple core assumptions. As such, we chose ordinal regression for the modelling in our main analyses.

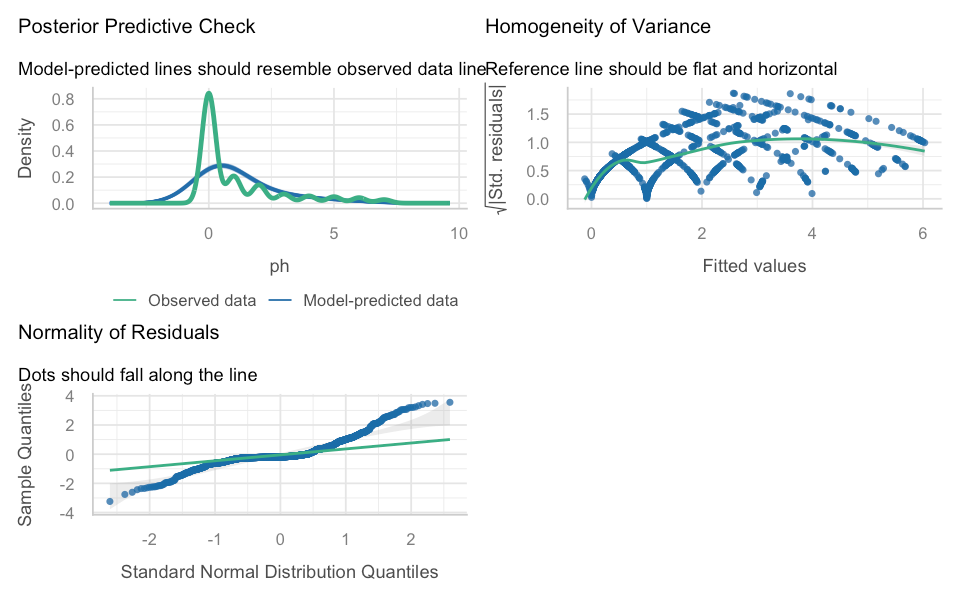

**Figure S1. Assumption checks of a linear mixed effect regression model of simulated data of PGS predicting autistic trait score.** Autistic trait score was non-normally distributed. Indeed, multiple assumptions were not met when using linear regression modelling.

#### Post hoc analyses and model fit

Estimated marginal means (EMMs) provide the predicted marginal effect for each level of the categorical variables of interest (rater and sex), while estimated marginal trends (EMTs) estimate the slopes for continuous predictor PGS separately (marginally) for these groups. We estimated EMMs and EMTs while holding all independent variables, not under direct investigation, constant at their mean (e.g. PGS = 0, the mean effect of genotyping platforms, etc.).

We calculated marginal and conditional R² values following Nakagawa et al.^14^ as implemented for *clmm* in the performance package^13^. For ordinal models, these are computed on the continuous latent scale, not on the observed ordinal outcome scale. Marginal R² represents the variance explained by fixed effects. Conditional R² represents variance explained by fixed + random effects.

### Measurement Invariance

When comparing sum scores across groups, it is essential that the items in the scale are not biased across these groups, a property known as measurement invariance (MI). To draw conclusions about sex differences in the summed autistic trait score, we need to establish that a sex difference reflects a difference in the latent variable, and not a difference in the items themselves. This is the case when the effect of sex on the items is entirely mediated through the latent variable (Figure S2). Typically, MI is tested with multi-group structural equation modelling (SEM), by fitting nested models with increasing equality constraints on the parameters across the groups of interest (in our case, sex). Below, we describe the modelling, evaluation, and results of the MI analysis of the So et al.^2^ ASEBA autism subscale per rater.

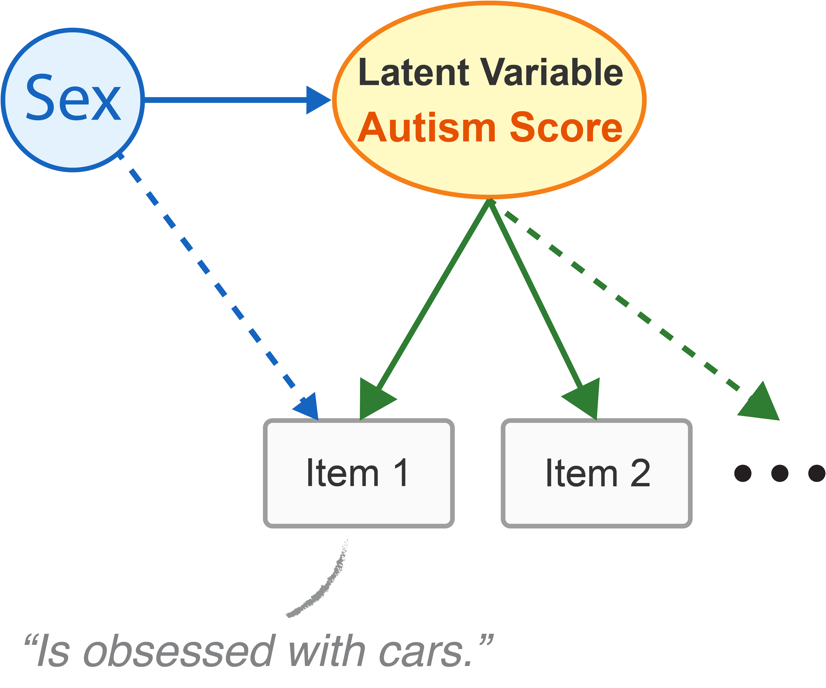

**Figure S2: Measurement Invariance example.** Presented is a hypothetical item “Is obsessed with cars” and a latent variable representing the autism sum score. The effect of sex on the item must be entirely mediated through the latent variable for the item to be considered measurement invariant with respect to sex.

#### Model specification and parameterisation

MI analyses were performed using Mplus Version 8^15^ by fitting single common factor confirmatory factor (CFA) models to the 9 (YSR) or 10 (CBCL & TRF) items of the autism scale. As the item responses are ordinal, we used ordinal factor modelling based on Mplus Theta parameterisation^16^, and the Weighted Least Squares Mean and Variance Adjusted (WLSMV) estimator. As participants are twins and their data are clustered in families, we added the Mplus correction for clustering. We fitted two-group CFA models with sex as the grouping variable with respect to which we investigated MI of the items.

The sequence of models includes increasingly restrictive constraints that can be statistically evaluated as the models are nested, as laid out in Table S1. We first fitted the configural model to establish that the single common factor model fits in the male and female samples. This model includes the standard scaling constraint, but does not include any equality constraints across sex. Next, we fitted the metric invariance model, in which the factor loadings are constrained to be equal across sex. Subsequently, we fitted the strong factorial invariance model, which includes the constraint that the factor loadings and item thresholds are equal across sex. In this model, the mean and variance of the common factor vary freely across sex. Next, we considered the strict factorial invariance model, which includes equal factor loadings, thresholds, and residual variances between the sexes. This represents complete MI, and sex differences are limited to differences in the means and variance of the common factor (i.e. the latent variable). To interpret sex differences in mean autistic trait scores as reflecting differences in the latent factor of autistic traits, strong or strict factorial invariance must be established, allowing latent factor means to be meaningfully compared across sex. Finally, a full invariance model is fit where additionally the latent factor mean is constrained. If at least the strong model shows an adequate fit, the full model can be compared to it. Here, rejection of the full model indicates a difference in group means of the latent variable; i.e. a meaningful difference in autistic trait score between the sexes after accounting for the measurement properties of the scale.

#### Model evaluation

Following van der Sluis et al.^17^, we evaluated the overall fit of the models using the Comparative Fit Index (CFI) and the root mean square error of approximation (RMSEA). For the CFI, a value of 0.95 or higher indicates a good model fit. For RMSEA, a value greater than 0.10 indicates a poor fit; a value between 0.05 and 0.08 indicates an adequate fit; and a value below 0.05 indicates a good fit.

The configural models were required to indicate a good fit before subsequent models were run. In case of a poor fit, the modification indices were checked for correlated residuals. An extended configural model was then made with specified covariance between the residuals of these items. Further nested models were then run with this specified covariance present.

We used $\chi^{2}$ difference tests to test the constraints associated with increasingly restrictive models, by comparing a given model in the sequence with the preceding model (configural vs. metric, metric vs. strong, strong vs. strict). For comparing models with the Weighted Least Squares Mean and Variance Adjusted (WLSMV) estimator, Mplus provides the DIFFTEST^15^ (section 13.12). The $p$-values of these tests are interpreted alongside the specific model fit indices and modification indices to evaluate the level of (non-)MI. Given our large sample size and high power for $\chi^{2}$ difference testing, we used $\alpha=.01$ for the DIFFTEST threshold. A significant DIFFTEST suggests that the equality constraints of a given model should be rejected. Finally, the full model was tested with DIFFTEST against the strong invariance model to evaluate group differences in the common factor mean.

A further step in investigating MI is investigating the modification indices of a model. These quantify the expected decrease in the model $\chi^{2}$ statistic if a currently fixed or constrained parameter (e.g. a cross-loading or residual covariance) were freely estimated. As the equality constraints refer to multiple parameters, we inspected the modification indices to determine the source of the model rejection. In the context of MI testing, modification indices are used diagnostically to identify specific equality constraints on an item level that contribute most to model misfit across groups. Rejection of a model in combination with high modification indices for a specific item indicates a sex bias for that item. For instance, in comparing the configural and the metric models, the DIFFTEST tests the equality of the factor loadings. If the metric invariance model were rejected, we would inspect the modification indices of the factor loadings to explore the reason for the rejection. We took a threshold value for any items with a modification index of 20 or greater to indicate a meaningful bias with respect to sex. Subsequently, a set of models was fitted without these items, and the model evaluation was performed again.

#### Measurement Invariance results show a sex bias in item 1

Configural models for Father, Teacher, and Self rater groups showed moderately inadequate CFI. Modification indices showed correlated residuals between items 42 ("Would rather be alone than with others") and 111 ("Withdrawn, doesn’t get involved with others"). These items ostensibly measure the same construct and were subsequently specified with covariance between the residuals in the extended model. Here, the CFI substantially improved over the base model above the good fit threshold. For completeness, the Mother model was also extended.

For the Self-extended model, metric invariance was not obtained following a $\chi^{2}$ difference test, indicating non-MI in factor loadings. For the Mother model and Teacher-extended model, the metric model showed a barely significant $\chi^{2}$ test result at our threshold ($\alpha=.01)$. For the Father-extended model, only strict invariance failed, indicating non-MI on the level of residual variance which we deemed acceptable.

Next, we inspected the modification indices of the rejected models. The Teacher-extended metric model and Father-extended strict model showed no modification indices above our threshold, indicating no substantial bias in individual items. In the Mother- and Self-extended metric models, item 1 ("Acts too young for his/her age") showed a sex bias. The expected parameter changes indicate that this item reflects the latent trait more strongly for boys. A sex bias according to MI means that boys score higher on this item as a function of sex rather than because of the latent trait of autism. Indeed, between the ages of 12–14, males and females show a differing development^18^, and it is logical to assume that this is reflected in the item.

All extended models were subsequently rerun, excluding item 1 (including Father and Teacher models for completeness). In the DIFFTEST, the Mother models now reached strong invariance, only failing at strict. Again, we found significant $\chi^{2}$ difference test results for the metric models of Teacher and Self. However, the fit indices improved substantially, and each nested model indicated good fit. Furthermore, no item in these models showed a bias above our threshold in the modification indices. As such, we accepted these models and deemed them sufficiently MI to investigate mean differences in the autistic trait score. Following these results, the main analyses of this study were rerun excluding item 1 as explained in the sensitivity analyses.

In summary, we identified the accepted models for each rater up to at least strong invariance as follows: Father; Mother-extended without item 1; Teacher-extended; Self-extended without item 1. Next, for all raters, we found a rejection of the full invariance model. This indicates a sex difference in their latent factor mean, reflecting a meaningful difference in their autistic trait score. The modification indices confirmed that for mother, father, and teacher raters, males scored higher. In contrast, for self-rated, females scored higher. These results are consistent with the results of the main analyses.

Full MI results can be found in Supplementary Tables S21–S22, including fit indices and modification indices for items of models with a significant $\chi^{2}$ difference test result. Full model outputs including modification indices can be found on GitHub.

### Sample descriptives

Histograms of age and date of assessment are shown in Figure S3 and Figure S4. Figures include the dataset for the sensitivity analysis where the YSR was taken in a sub population via self-report at age 12. Notably for date of assessment, the self-reports showed a substantially smaller range. For the YSR at age 12, the reports were taken at an earlier date. For age, the self-reports showed a substantially higher mean. The YSR at age 12 showed similar distribution to the other raters.

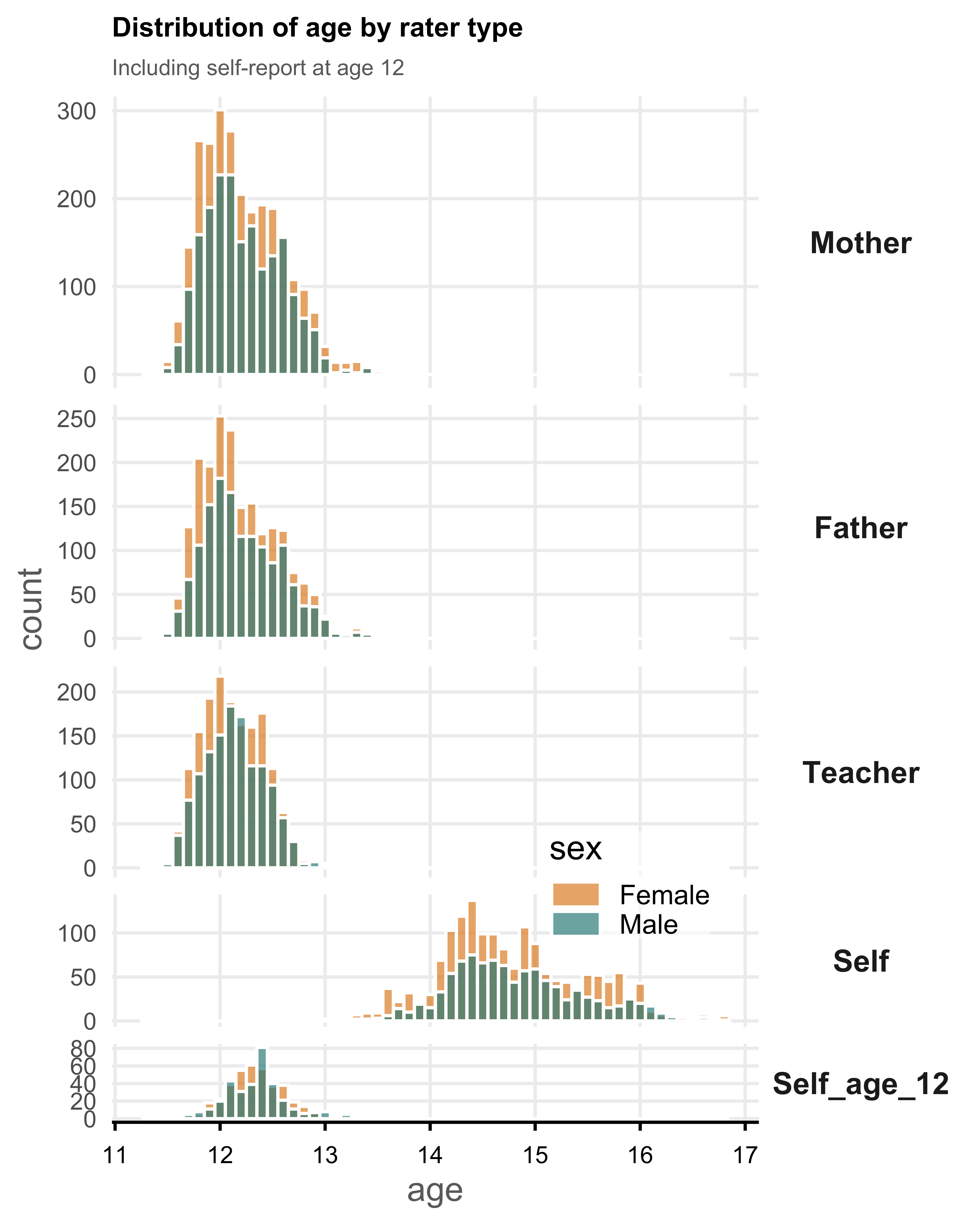

**Figure S3: Histogram of age separated by rater and sex.** Self at age 12 reflects the sensitivity analysis sub-population of participants for which the YSR was taken at age 12.

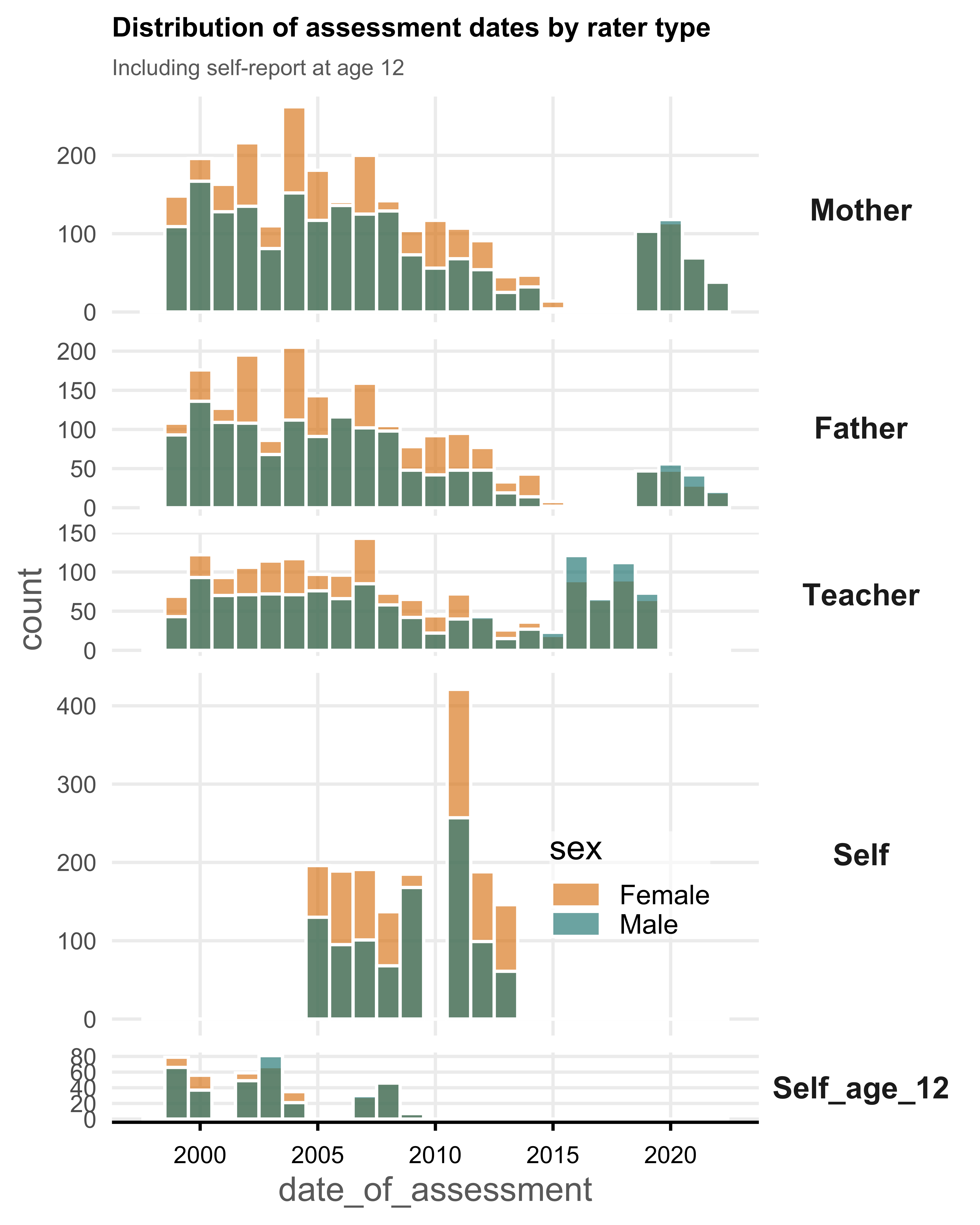

**Figure S4:** **Histogram of assessment date separated by rater and sex.** Self at age 12 reflects the sensitivity analysis sub-population of participants for which the YSR was taken at age 12.

### Sensitivity analyses

Item 80 is not available in the YSR questionnaire. Additionally, measurement invariance suggested a bias in item 1. As such, we performed a sensitivity analysis by removing these items and rerunning the models. Results of the analysis did not differ from our main results (Supplementary Tables S23–S26).

Because not all raters were present for each participant, rater effects could be due to differences in sample composition. To address this, we ran the models in a subsample of individuals with all raters present. Due to a substantially reduced sample (male *n* = 346, female *n* = 556), power was limited. The previously significant effect of females scoring higher in the self-rated group no longer reached significance (*OR* = 0.81, 95% *CI* [0.58, 1.13], *p* = .214). However, the overall pattern of results is comparable, and the main conclusions remain unchanged (Supplementary Tables S27–S30).

We investigated whether sex differences across raters persist at a high level of PGS. We compared males and females across raters using estimated marginal effects at a PGS of 2 *SD* above the mean (see Supplementary Table S31). Results showed a similar pattern of external raters rating males higher, and females showing higher self-ratings.

We compared model 5 (Supplementary Table S11, M5 in main text) to the main results as a sensitivity analysis. In general, results stayed the same, despite substantial increase in the number of modelled parameters. Notably however, self-rated scores were no longer higher for females (*OR* = 1.20, 95% *CI* [0.57, 2.53], *p* = .626), and fathers no longer rated males higher (*OR* = 1.36, 95% *CI* [0.89, 2.07], *p* = .207). Further investigation revealed substantial multicollinearity between main effects and covariate interactions introduced in model 5. Specifically, age, rater, and sex showed high correlations with their interactions. To further investigate this, we modelled only the interaction of rater × sex in the self-rated subset specifically with interactions of age × sex and assessment date × sex as covariates. A large multicollinearity was present between sex and age × sex. Crucially, we rescaled age and date within this data subset. The high correlation disappeared after rescaling age, and the effect of sex on autistic trait score level again showed higher scores for females (*OR* = 0.69, 95% *CI* [0.56, 0.84], *p* < .001). We applied this technique to model 5 by rescaling age and date within each rater separately. Indeed, the result of fathers rating males higher returned (*OR* = 1.54, 95% *CI* [1.08, 2.20], *p* = .022), but self-rated scores were still not significantly higher for females (*OR* = 0.69, 95% *CI* [0.44, 1.10], *p* = .117). Results indicate that age differences between the self-rated and external-rated introduced multicollinearity between sex and the covariate interactions^19^.

The mean age at rating in the self-rated group is substantially higher than in the other raters (Table 1 in main text). As such, in our results we could not reliably distinguish between age and rater effects. To further investigate age effects, we performed a sensitivity analysis in a YNTR subsample with self-report data available at age 12 (male *n* = 337, mean age = 12.3, *SD* 0.29; female *n* = 382, mean age = 12.4, *SD* 0.26). Here, we modelled sex, assessment date, and age predicting autistic trait score (we only tested rater × sex). Contrary to main analysis results, no sex effect was found (*OR* = 1.03, 95% *CI* [0.69, 1.55], *p* = .881), indicating similar autistic trait scores between males and females. However, the 95% *CI* are large, likely attributable to the small sample size. These results, combined with model 5 results indicate we cannot categorically distinguish age and rater effects.

### Sex differences and sex amplification under the Liability Threshold Model

Two interpretations exist under the Liability Threshold Model (LTM) that could lead to more male autism diagnoses^20,21^. The LTM posits an underlying continuous liability distribution that results in a case status, such as an autism diagnosis, once a threshold is reached. Both biological and environmental factors jointly contribute to this liability.

First, the genetic sex amplification shown in Figure S5 assumes that autism-associated genetic variants systematically have a higher effect in males, leading to more males surpassing the threshold^22^. At an equal number of variants females would then show lower autistic trait levels, and more variants are needed to cross the threshold; the association between PGS and autistic traits would thus be weaker in females. Under this interpretation of the LTM, no main effect of sex needs to be present as indicated by a mean shift in liability or a dual-threshold. As shown in Figure S5, sex amplification would mean a higher male genetic variability. Indeed, Sandin et al.^21^ reported a higher family-based heritability of autism diagnosis in males, which they interpret as potentially reflecting greater male genetic variance. Sex amplification is one explanation of a higher male variability, but other sex-specific effects could be present such as via different variants as reflected by recent findings of a genetic correlation between the sexes being significantly different from 1 (*r_g_* ≈ 0.8)^23^. Of note, a difference in liability between the sexes does not necessitate a difference in genetic variability such as via genetic sex amplification, and could be due to other biological factors, or could reflect environmental effects.

Second is the dual-threshold model shown in Figure S6, where the sexes have a differing threshold under the LTM, resulting in more male cases. Another formulation of this model is one threshold, but a mean shift in liability between the sexes, such that females have a lower mean liability^20^. A higher threshold for females might arise from a necessity for more extreme traits before these are recognised in their environment, which would indicate societal barriers and/or diagnostic biases, including rater effects. Under the dual-threshold interpretation, the variance of the liability is the same for both sexes, but females would have a higher threshold, implying they need a higher liability before they reach the threshold. In terms of genetics this could mean a higher genetic liability such as a higher polygenic load. Indeed, in a previous systematic review we found that autistic females without an intellectual disability showed a higher mean PGS^24^. This higher threshold (and the higher mean PGS) could be due to biological factors causing females to develop fewer autistic traits or could reflect societal barriers and diagnostic biases where females need more autistic traits before these are recognised.

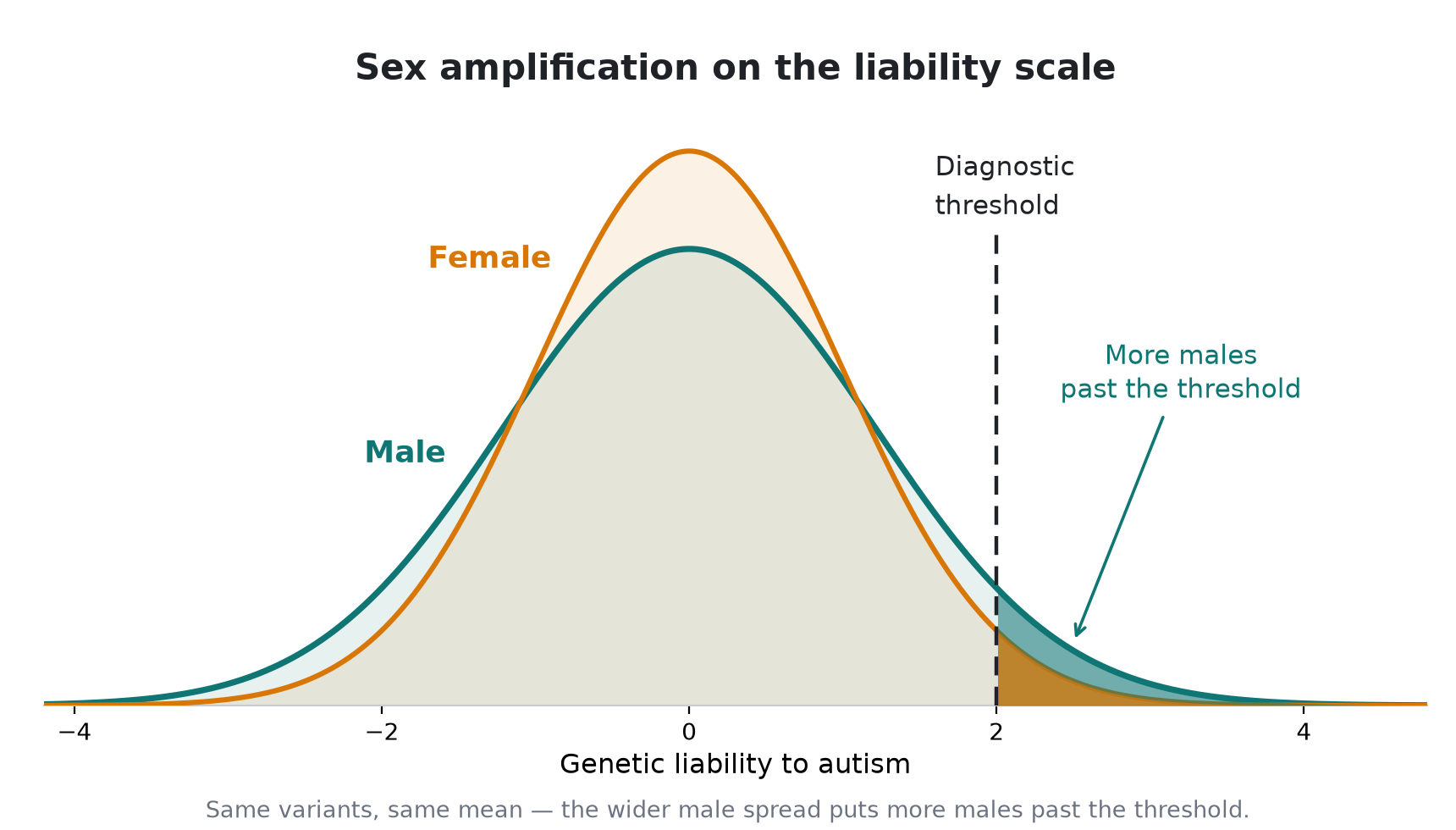

**Figure S5. Sex amplification in autism genetics.** The figure shows genetic effects on the liability scale. A greater male genetic variance leads to a higher number of males past the diagnostic threshold. In terms of polygenic scores, a higher association with the autism phenotype for males would suggest a sex amplification effect.

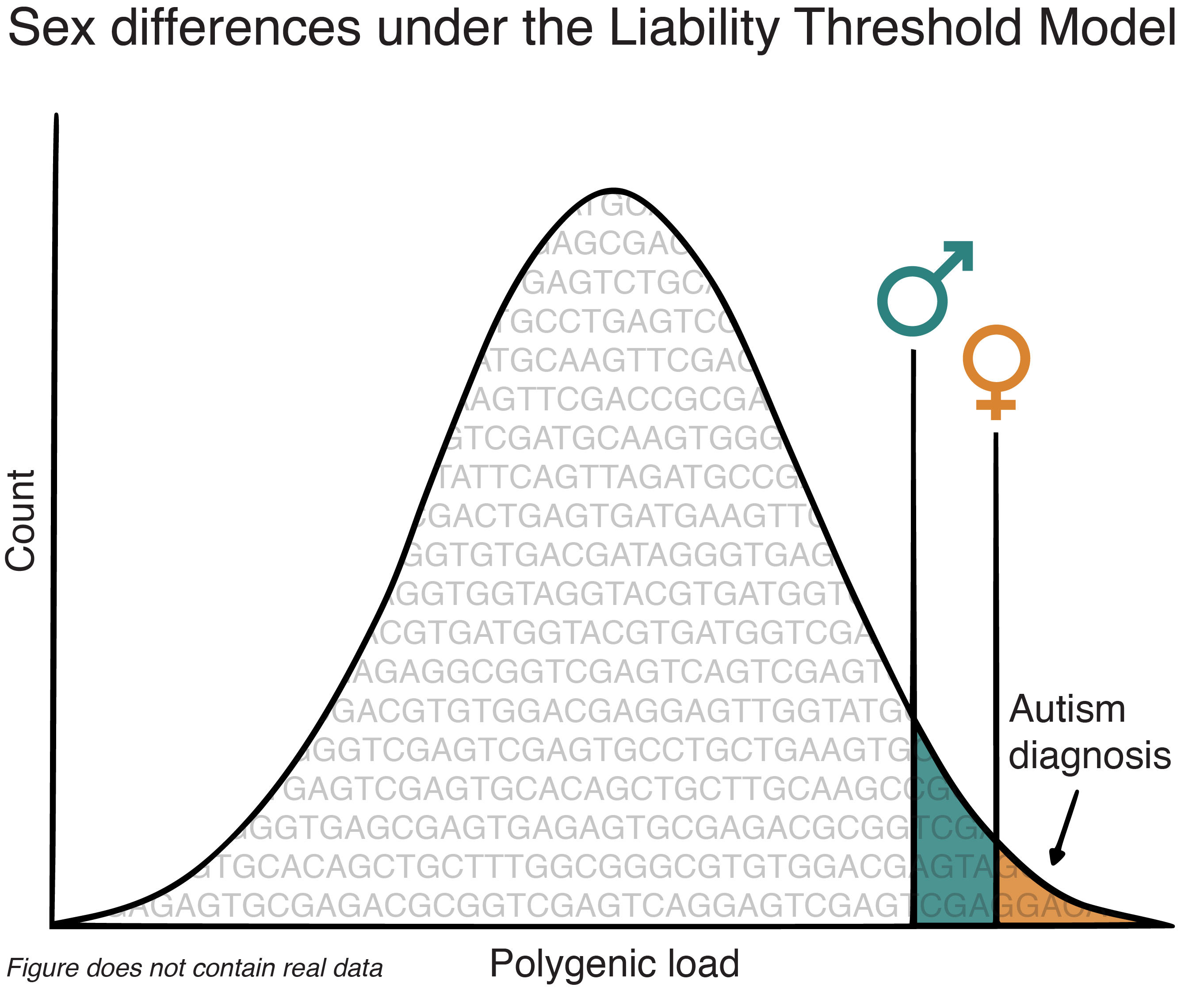

**Figure S6: Sex differences in polygenic load under the dual-threshold Liability Threshold Model (LTM).** Previous work by our group^24^ found a higher mean polygenic scores in autistic females without an intellectual disability. Under the LTM this suggest a higher threshold to reach an autism diagnosis. A sex difference in threshold can reflect biological differences in trait expression or environmental/societal barriers.

### References

1.	Fyfe, C. *et al.* Time trends in the male to female ratio for autism incidence: population based, prospectively collected, birth cohort study. *BMJ* **392**, e084164 (2026).

2.	So, P. *et al.* Using the Child Behavior Checklist and the Teacher’s Report Form for identification of children with autism spectrum disorders. *Autism* **17**, 595–607 (2013).

3.	Browning, B. L., Zhou, Y. & Browning, S. R. A One-Penny Imputed Genome from Next-Generation Reference Panels. *Am. J. Hum. Genet.* **103**, 338–348 (2018).

4.	Fairley, S., Lowy-Gallego, E., Perry, E. & Flicek, P. The International Genome Sample Resource (IGSR) collection of open human genomic variation resources. *Nucleic Acids Res.* **48**, D941–D947 (2020).

5.	Danecek, P. *et al.* Twelve years of samtools and bcftools. *GigaScience* **10**, giab008 (2021).

6.	Chang, C. C. *et al.* Second-generation PLINK: rising to the challenge of larger and richer datasets. *GigaScience* **4**, s13742-015-0047–8 (2015).

7.	Purcell, S. & Chang, C. PLINK1.9. (2015).

8.	Abdellaoui, A. *et al.* Population structure, migration, and diversifying selection in the Netherlands. *Eur. J. Hum. Genet.* **21**, 1277–1285 (2013).

9.	Patterson, N., Price, A. L. & Reich, D. Population Structure and Eigenanalysis. *PLoS Genet.* **2**, e190 (2006).

10.	Zeng, J. *et al.* Signatures of negative selection in the genetic architecture of human complex traits. *Nat. Genet.* **50**, 746–753 (2018).

11.	Zheng, Z. *et al.* Leveraging functional genomic annotations and genome coverage to improve polygenic prediction of complex traits within and between ancestries. *Nat. Genet.* **56**, 767–777 (2024).

12.	Hoaglin, D. C., Iglewicz, B. & Tukey, J. W. Performance of Some Resistant Rules for Outlier Labeling. *J. Am. Stat. Assoc.* **81**, 991–999 (1986).

13.	Lüdecke, D., Ben-Shachar, M. S., Patil, I., Waggoner, P. & Makowski, D. performance: An R Package for Assessment, Comparison and Testing of Statistical Models. *J. Open Source Softw.* **6**, 3139 (2021).

14.	Nakagawa, S., Johnson, P. C. D. & Schielzeth, H. The coefficient of determination R2 and intra-class correlation coefficient from generalized linear mixed-effects models revisited and expanded. *J. R. Soc. Interface* **14**, 20170213 (2017).

15.	Muthén, L. K. & Muthén, B. *Mplus User’s Guide*. (Los Angeles, CA: Muthén & Muthén, 2017).

16.	Millsap, R. E. & Yun-Tein, J. Assessing Factorial Invariance in Ordered-Categorical Measures. *Multivariate Behav. Res.* **39**, 479–515 (2004).

17.	van der Sluis, S. *et al.* Sex differences and gender‐invariance of mother‐reported childhood problem behavior. *Int. J. Methods Psychiatr. Res.* **26**, e1498 (2017).

18.	Marceau, K., Ram, N., Houts, R. M., Grimm, K. J. & Susman, E. J. Individual Differences in Boys’ and Girls’ Timing and Tempo of Puberty: Modeling Development With Nonlinear Growth Models. *Dev. Psychol.* **47**, 1389–1409 (2011).

19.	Keller, M. C. Gene-by-environment interaction studies have not properly controlled for potential confounders: The problem and the (simple) solution. *Biol. Psychiatry* **75**, 18–24 (2014).

20.	Dougherty, J. D. *et al.* Can the “female protective effect” liability threshold model explain sex differences in autism spectrum disorder? *Neuron* **110**, 3243–3262 (2022).

21.	Sandin, S. *et al.* Examining Sex Differences in Autism Heritability. *JAMA Psychiatry* **81**, 673–680 (2024).

22.	Zhu, C. *et al.* Amplification is the primary mode of gene-by-sex interaction in complex human traits. *Cell Genom.* **3**, 100297 (2023).

23.	Zhang, X. *et al.* Polygenic and developmental profiles of autism differ by age at diagnosis. *Nature* **646**, 1146–1155 (2025).

24.	Sollie, T., Akingbuwa, W. A., de Wit, M. M., Badura, A. & Polderman, T. J. C. Sex Differences in the role of Additive Genetic Variants in Autism: A Systematic Review. *medRxiv* 2025.12.23.25342905 (2025) doi:10.64898/2025.12.23.25342905.
